# Implication of DNA methylation changes at chromosome 1q21.1 in the brain pathology of Primary Progressive Multiple Sclerosis

**DOI:** 10.1101/2022.05.06.22274611

**Authors:** Majid Pahlevan Kakhki, Chiara Starvaggi Cucuzza, Antonino Giordano, Tejaswi Venkata S. Badam, Pernilla Strid, Klementy Shchetynsky, Adil Harroud, Alexandra Gyllenberg, Yun Liu, Sanjaykumar Boddul, Tojo James, Melissa Sorosina, Massimo Filippi, Federica Esposito, Fredrik Wermeling, Mika Gustafsson, Patrizia Casaccia, Ingrid Kockum, Jan Hillert, Tomas Olsson, Lara Kular, Maja Jagodic

## Abstract

Multiple Sclerosis (MS) is a heterogeneous inflammatory and neurodegenerative disease of the central nervous system with an unpredictable course toward progressive disability. Understanding and treating progressive MS remains extremely challenging due to the limited knowledge of the underlying mechanisms. We examined the molecular changes associated with primary progressive MS (PPMS) using a cross-tissue (blood and post-mortem brain) and multilayered data (genetic, epigenetic, transcriptomic) from independent cohorts. We identified and replicated hypermethylation of an intergenic region within the chromosome 1q21.1 locus in the blood of PPMS patients compared to other MS patients and healthy individuals. We next revealed that methylation is under genetic control both in the blood and brain. Genetic analysis in the largest to date PPMS dataset yielded evidence of association of genetic variations in the 1q21.1 locus with PPMS risk. Several variants affected both 1q21.1 methylation and the expression of proximal genes (*CHD1L, PRKAB2, FMO5*) in the brain, suggesting a genetic-epigenetic-transcriptional interplay in PPMS pathogenesis. We addressed the causal link between methylation and expression using reporter systems and dCas9-TET1-induced CpG demethylation in the 1q21.1 region, which resulted in upregulation of *CHD1L* and *PRKAB2* genes in SH-SY5Y neuron-like cells. Independent exploration using unbiased correlation network analysis confirmed the putative implication of *CHD1L* and *PRKAB2* in brain processes in PPMS patients. Thus, several lines of evidence suggest that distinct molecular changes in 1q21.1 locus, known to be important for brain development and disorders, associate with genetic predisposition to high methylation in PPMS patients that regulates the expression of proximal genes.

**Significance Statement:** Multiple sclerosis (MS) is a long-lasting neurological disease affecting young individuals that occurs when the body’s natural guard (immune system) attacks the brain cells. There are currently no efficient treatments for the progressive form of MS disease, probably because the mechanisms behind MS progression are still largely unknown. Thus, treatment of progressive MS remains the greatest challenge in managing patients. We aim to tackle this issue using the emerging field called “epigenetics” which has the potential to explain the impact of genetic and environmental risk factors in MS. In this project, by using unique clinical material and novel epigenetic tools, we identified new mechanisms involved in MS progression and putative candidates for targeted epigenetic therapy of progressive MS patients.

## Introduction

Multiple Sclerosis (MS) is an inflammatory disease of the central nervous system (CNS) affecting young adults and leading to unpredictable and progressive physical and cognitive impairments. The pathological hallmarks of MS are represented by focal plaques of primary demyelination and di□use neurodegeneration in the grey and white matter of the brain and spinal cord (1). The most common form of disease, relapsing-remitting MS (RRMS), affects twice as many women as men and presents itself with neurological relapses followed by periods of partial or complete remission. In most cases, this inflammatory stage is followed by a secondary progressive phase of MS (SPMS) after several years. About 10-15% of patients, manifest a primary progressive form of MS disease (PPMS) with uninterrupted progression starting from disease onset, although relapses may occur (2). Beyond epidemiological differences, both progressive forms of MS share similar features that differ from RRMS stage, notably a later onset (40 years vs. 30 years in RRMS) and a prevalence of CNS-intrinsic inflammatory and degenerative pathological processes as opposed to the predominant role of peripheral immune influence in RRMS stage (3). However, while considerable progress has been achieved in deciphering the genetic variation predisposing for disease, the mechanisms underpinning disease progression and severity remain unresolved. Importantly, conventional immunomodulatory therapies are ineffective in progressive patients (4, 5), reinforcing the need to better clarify processes underlying MS courses.

Disease trajectory likely relies on a complex interplay between genetic and non-genetic factors (6). Epigenetic modifications such as DNA methylation, intersecting internal and external influences might provide novel opportunity to study the mechanisms underlying the different forms of MS disease. DNA methylation, which regulates gene expression without altering the genetic code, is the most studied epigenetic mark and its implications in MS pathogenesis have been reported in immune and nervous cells of MS patients (7-12). We have shown that the major risk variant *HLA-DRB1*15:01* may mediate risk for MS via changes in *HLA-DRB1* DNA methylation and subsequent expression (11). Recent comparative whole-genome DNA methylation studies shed further light on molecular mechanisms behind MS pathogenesis (9, 13), the vast majority of them focusing on RRMS stage of disease.

In this study, we aimed to examine the molecular changes that associate with MS course. Using a multilayered (genetic, epigenetic, transcriptomic) and cross-tissue (blood, brain) approach in cohorts of MS patients and controls, we demonstrate an interplay between regional genetic variation, DNA methylation and gene expression in the chromosome 1q21.1 region, in PPMS patients specifically. We addressed the link between these three molecular layers using in-vitro reporter gene assays and CRISPR/dCas9 epigenome editing. Our findings suggest that genetically controlled methylation at 1q21.1 locus might contribute to PPMS pathology though transcriptional regulation of proximal genes, particularly *CHD1L*.

## Results

### Hypermethylation of an intergenic region in the 1q21.1 locus of PPMS patients

We profiled genome-wide DNA methylation in the blood of RRMS, SPMS and rare PPMS patients and matched healthy controls (HC, cohort 1, n = 279, Fig. 1, SI Appendix Table S1), using the Illumina Infinium HumanMethylation450 BeadChip (450K array). After adjustment for confounders, we identified one differentially methylated region (DMR) in PPMS patients in comparison with HC (*P* = 6.16 × 10^−6^), RRMS (*P* = 2.51 × 10^−6^) and SPMS (*P* = 7.33 × 10^−6^) patients (FWER < 0.05, Fig. 2a, SI Appendix Table S2). The DMR, located in an intergenic CG-rich region on chromosome 1 (q21.1): 146549909–146551201 (GRCh37/hg19), consists of 8 consecutive CpG probes displaying hypermethylation in PPMS patients compared to RRMS, SPMS as well as HC (Fig. 2b). Considering the low number of PPMS patients (n = 4), we selected an independent cohort for replication including 36 PPMS and 48 RRMS patients, matched for gender, age and disease duration (cohort 2, Fig. 1, SI Appendix Table S1). We designed pyrosequencing assays covering seven CpG sites (CpG 1-7), including the 450K probes exhibiting the largest changes cg21263664 (CpG 1), cg03526459 (CpG 3), cg16814483 (CpG 5), cg02487331 (CpG 7) and three additional adjacent CpG sites located between theses 450K-CpGs (Fig. 2a). A total of six out of these seven CpGs were found significantly hypermethylated in PPMS compared to RRMS patients (Fig. 2c). These data imply that PPMS patients display hypermethylation of an intergenic region within the 1q21.1 locus, suggesting a potential involvement in the PPMS pathogenesis.

**Figure 1.**
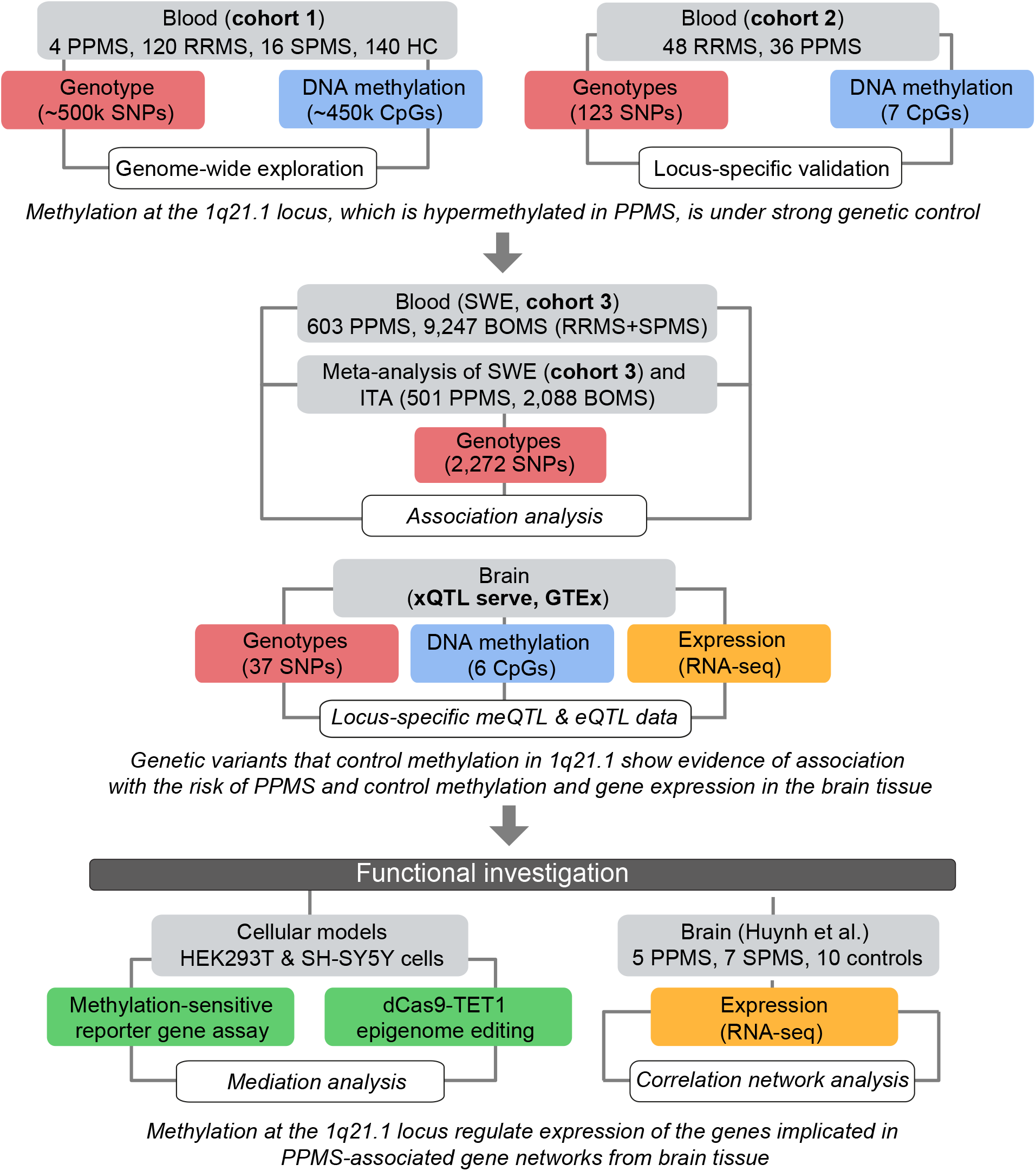
Workflow of the study. MS, multiple sclerosis; PPMS, primary progressive MS; SPMS, secondary progressive MS; RRMS, relapsing-remitting MS; SNP, single nucleotide polymorphism, meQTL and eQTL, methylation and expression quantitative trait loci. SWE, Swedish cohort; ITA, Italian cohort; META, meta-analysis of the SWE and ITA cohorts.

**Figure 2.**
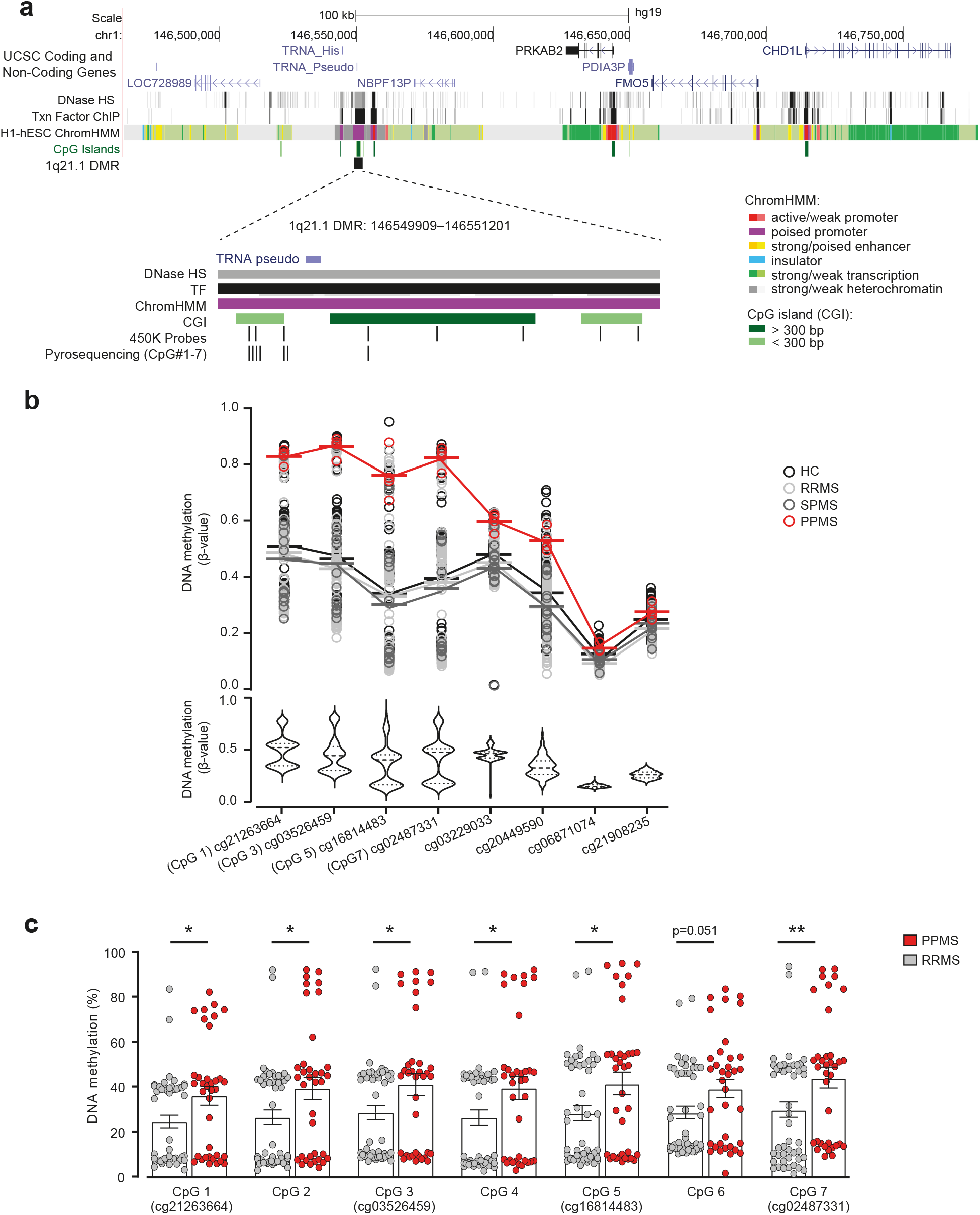
Hypermethylation of the 1q21.1 locus in primary progressive multiple sclerosis patients. **a**. Genomic annotation of the identified differentially methylated region (DMR) in 1q21.1, including 450K and pyrosequenced CpG probes and regulatory features, i.e. CpG island (CGI) and chromatin state segmentation by hidden Markov model (ChromHMM) from ENCODE/Broad. **b**. Dot plot and violin plot of DNA methylation differences at the identified DMR on chromosome 1: 146549909-146551201 (hg19) in PPMS (n = 4) compared to RRMS (n = 120), SPMS (n = 16) and HC (n = 139), obtained using Illumina 450K in cohort 1. **c**. Replication of methylation at 7 CpGs (CpG 1-7, including four 450K probes) in an independent cohort (RRMS n = 48, PPMS n = 36) using pyrosequencing. * and ** correspond to P-value < 0.5 and 0.01, respectively, using non-parametric Mann-Whitney U test. MS, multiple sclerosis; PPMS, primary progressive MS; RRMS, relapsing-remitting MS; SPMS, secondary progressive MS; HC, healthy controls; DNase HS, DNase I hypersensitive sites; Txn factor ChIP, Transcription Factor chromatin immunoprecipitation.

### Methylation of the PPMS-associated 1q21.1 locus is under strong genetic control in the blood and the CNS

Previous studies have revealed a complex interplay between genetic and epigenetic variations by reporting that part of the epigenome is under genetic control in general (14, 15), and more particularly, that genetically dependent DNA methylation can mediate risk to develop MS (11). Interestingly, our data revealed that methylation clustered within three methylation levels (i.e. low, medium and high) in both cohorts (Fig. 2b-c), a pattern typically observed for genetically controlled methylation. To test for potential genetic dependence, we performed genome-wide methylation Quantitative Trait Loci (meQTL) analysis in cohort 1 and found 19 SNPs, located in the extended 1q21.1 locus (−50 kb to +400 kb), associating with methylation levels of the DMR, with strongest effects coming from rs1969869, rs21327, rs647596, rs10900384 and rs12401360 (genotype *vs*. methylation estimate = 0.19, p < 10^−16^) (Fig. 3a, SI Appendix Table S3). To further validate this finding, we conducted locus-specific meQTL analysis in cohort 2 by testing all SNPs covering ± 500 kb window encompassing the DMR against each pyrosequenced CpG. Of note, only eight out of the 19 meQTL-SNPs identified in cohort 1 could be assessed in this cohort. Out of the 123 SNPs tested in cohort 2, two variants showed significant (rs1969869, rs4950357, p < 10^−8^) and two suggestive (rs647596, rs10900384, p < 10^−5^) associations with CpGs methylation (Fig. 3a). An example of genetic control of methylation at CpG 4 (pyrosequenced CpG located between cg03526459 and cg16814483, Fig. 2c) is illustrated in Figure 3b. The two strongest SNPs, rs1969869 and rs4950357, displayed positive association between the minor allele and CpG methylation levels (Fig. 3a, SI Appendix Table S4). In addition to these four SNPs, 66 SNPs demonstrated nominal association with methylation (p < 0.05) (SI Appendix Table S4). Overall, all eight overlapping SNPs identified in cohort 1 displayed significant and similar direction of changes also in the cohort 2 (Fig. 3a, SI Appendix Tables S3 and S4). Many of the SNPs with similar effect, i.e. minor allele associating with either high or low methylation, were in strong linkage disequilibrium (LD > 0.8) and were therefore grouped into LD clusters (Fig. 3a).

**Figure 3.**
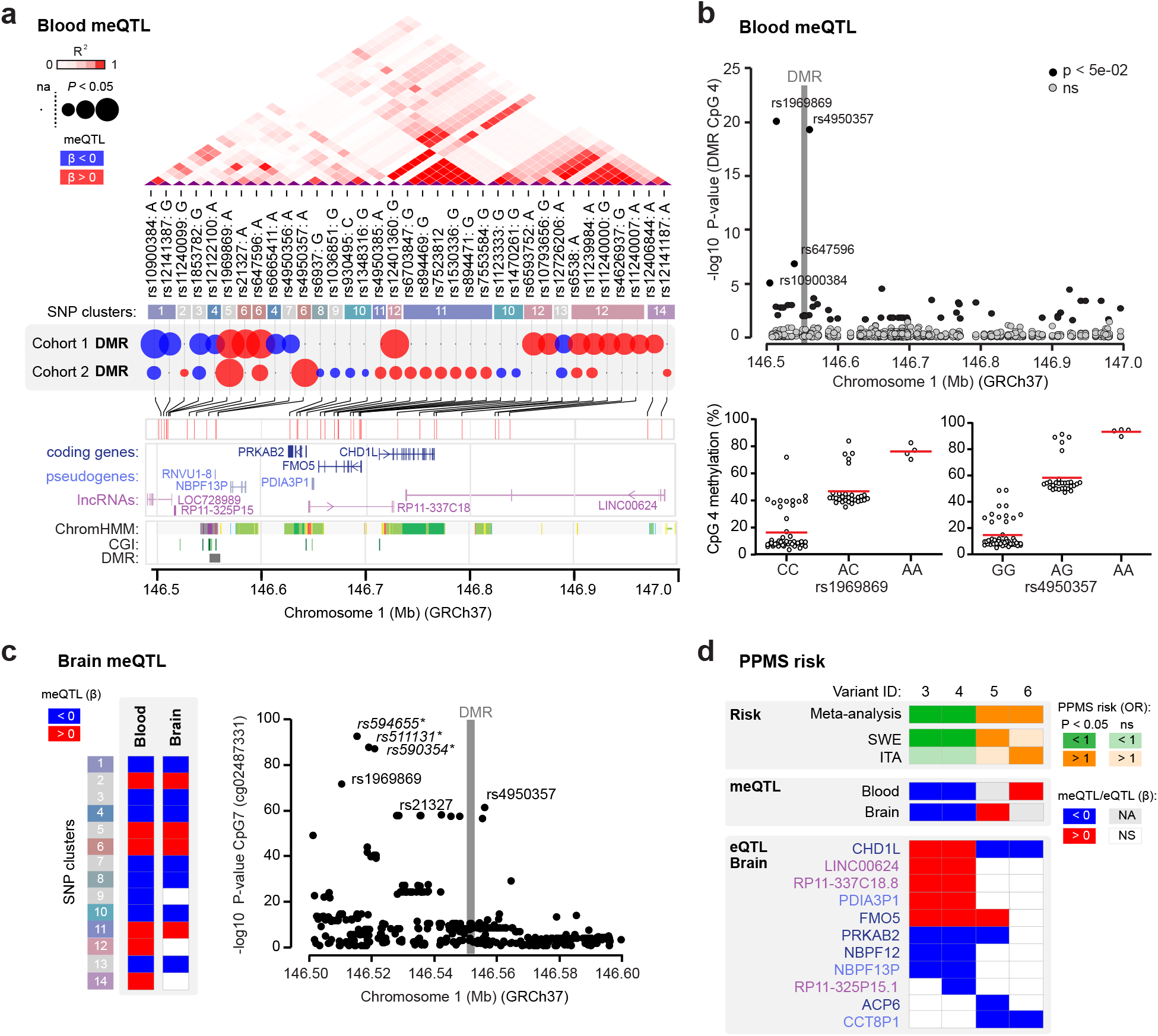
Genetic control of methylation and gene expression in the 1q21.1 locus. **a**. Genomic and functional annotation of the methylation-controlling variants (SNPs). Linkage disequilibrium (LD) is indicated by shades of red (increasing R^2^ from light to dark red) and clustering of SNPs is based on high LD between SNPs (R^2^ > 0.8). The effects of SNPs on DMR methylation and PPMS risk are depicted using colored circles: the size and color of circles represent significance (–log10 P-value) and direction of the effect, respectively, conferred by the minor allele. Gene location and regulatory features, i.e. CpG island (CGI) and chromatin state segmentation by hidden Markov model (ChromHMM), from ENCODE/Broad. **b**. Association between genetic variation at the extended locus (upper panel) and DNA methylation in blood at CpG 4 of the 1q21.1 DMR, obtained using meQTL analysis in cohort 2 (n=82). Significance is represented as –log10 (P-value). Association of the two strongest variants rs1969869 and rs4950357 with CpG 4 (as an example) are depicted in the lower panel. **c**. Effects of SNPs within each SNP cluster on DNA methylation at 1q21.1 DMR in the brain (using xQTL serve platform (15)) with only significant associations are displayed in colors, blue and red colors representing negative and positive effect of the minor allele, respectively (left panel) and association between genetic variation in the extended locus and DNA methylation at cg02487331 (CpG7) (right panel). The location and code for the CpG numbers is presented in Figure 2c. Significance is represented as –log10 (P-value). *Indicates that the SNPs are in strong LD with rs21327 (LD = 0.7). **d**. Association between genetic variation, DNA methylation (using xQTL serve platform (15)) and gene expression (using GTEx database) in the extended locus for SNPs tagging four variants displaying significant association with PPMS in the meta-analysis of Swedish (SWE) and Italian (ITA) cohorts. Protective and risk genetic effects are depicted in green and orange, respectively. Green and orange colors reflect protective and risk effect on PPMS risk conferred by the genetic variants. Blue and red colors represent negative and positive effect of the minor allele, respectively, on methylation or expression. The data shown in this figure are available in SI Appendix Tables.

Our results implying a link between genetic variation and blood DNA methylation at the 1q21.1 region, a locus reported to be crucial for brain size (16, 17) and neuropsychiatric disorders (18-20), posed the question whether such effect might occur in the CNS as well. To address this, we examined putative genetic influences on the methylation at 1q21.1 DMR in the brain using publicly available meQTL data from brain tissue of 543 individuals (15). Results showed that SNPs corresponding to 11 out of 14 SNP clusters demonstrated strong meQTLs in the CNS as well, with similar direction of the effect as observed in the blood (Fig. 3c, SI Appendix Table S5), exemplified for cg02487331 (Fig. 3c). Altogether, these findings strongly suggest that methylation at CpGs in the 1q21.1 DMR are under genetic control in the blood and CNS compartments.

### Genetic variants in 1q21.1 predispose for PPMS and associate with methylation and gene expression changes in the CNS

We next asked whether the genetic variation in 1q21.1 also affects the risk of developing PPMS. We tested the association between all the imputed SNPs in the extended locus (3,057 SNPs, chr.1: 145695987-147526765, hg19) and disease course, comparing PPMS and bout-onset MS (BOMS including RRMS and SPMS) patients in a combined Swedish cohort of 9,850 patients (PPMS n = 603, BOMS n = 9,247) (cohort 3, Fig. 1, SI Appendix Table S1). A total of 574 SNPs showed evidence for nominal association (p < 0.05) with PPMS (SI Appendix Table S6). Globally, these SNPs mapped to 57 of the 69 identified LD blocks. Of note, 13 SNPs belonging to seven different LD blocks remained significant after Bonferroni correction for multiple testing (p < 7.2 × 10^−4^).

Brain-meQTL data were available for 149/574 SNPs, belonging to 13 LD blocks. The vast majority of them (147/149) showed evidence for meQTLs in the brain. All the protective variants were found to be associated with lower methylation at 1q21.1 DMR, while 7/8 risk variants for PPMS were associated with higher levels of methylation at 1q21.1 DMR in the brain (SI Appendix Table S7). To further reinforce the evidence of a genetic influence at 1q21.1 on the risk of developing PPMS, we conducted a meta-analysis between the aforementioned Swedish cohort and an Italian cohort of PPMS (n = 501) and BOMS (n = 2,088) individuals (2,272 SNPs). A total of 64 SNPs, from 16 different LD blocks, showed a nominal association (p < 0.05) and same direction of effect in both cohorts (Table 1, SI Appendix Table S8). Five SNPs, tagging two LD blocks, remained significant after correction for multiple testing accounting for LD blocks (p < 7.2 × 10^−4^) (Table 1). All the five SNPs were associated with an increased risk of PPMS, and annotation to brain-meQTL data confirmed that they also lead to higher methylation (Fig. 3d, SI Appendix Table S7). These findings suggest a functional link between high methylation at 1q21.1 DMR and the genetic predisposition to develop PPMS.

**Table 1.** A meta-analysis of genetic association studies with PPMS (n = 1,104) in comparison to BOMS (n = 11,335).

| SNP<sup>a</sup> | BP | CHR | LD# | Meta-analysis |  |  | SWE (n = 9,850) |  | ITA (n = 2,589) |  |
| --- | --- | --- | --- | --- | --- | --- | --- | --- | --- | --- |
|  |  |  |  | A1 | P | OR | P | OR | P | OR |
| rs900347 | 145726727 | 1 | 1 | A | 0.0285 | 0.90 | 0.0352 | 0.87 | 0.3624 | 0.94 |
| rs11800992 | 146501783 | 1 | 2 | A | 0.0093 | 0.82 | 0.0260 | 0.78 | 0.1399 | 0.85 |
| rs72691008 | 146708291 | 1 | 3 | A | 0.0017 | 0.69 | 0.0083 | 0.69 | 0.0895 | 0.70 |
| rs7514808 | 146570076 | 1 | 4 | A | 0.0286 | 0.89 | 0.0367 | 0.86 | 0.3632 | 0.93 |
| rs72706463 | 146615555 | 1 | 5 | A | 0.0458 | 1.29 | 0.1141 | 1.37 | 0.2002 | 1.24 |
| rs10453880 | 146693689 | 1 | 6 | T | 0.0065 | 1.26 | 0.1083 | 1.21 | 0.0236 | 1.32 |
| rs145465008 | 146630173 | 1 | 7 | C | 0.0293 | 1.51 | 0.4867 | 1.17 | 0.0048 | 2.54 |
| **rs115153075** | 146978424 | 1 | 8 | T | 0.0006 | 1.48 | 0.0058 | 1.42 | 0.0303 | 1.74 |
| **rs12096043** | 147060953 | 1 | 9 | T | 0.0001 | 1.84 | 0.0005 | 2.03 | 0.0633 | 1.58 |
| rs672619 | 147058575 | 1 | 10 | T | 0.0013 | 1.34 | 0.0374 | 1.32 | 0.0146 | 1.35 |
| rs10494248 | 147108260 | 1 | 11 | A | 0.0079 | 1.72 | 0.0270 | 1.68 | 0.1386 | 1.82 |
| rs140566115 | 147126437 | 1 | 12 | A | 0.0244 | 1.53 | 0.0335 | 1.66 | 0.3586 | 1.34 |
| rs116430961 | 147312516 | 1 | 13 | C | 0.0101 | 1.62 | 0.1335 | 1.44 | 0.0265 | 1.93 |
| rs61740912 | 147313970 | 1 | 14 | G | 0.0012 | 1.86 | 0.0098 | 1.81 | 0.0483 | 1.97 |
| rs112044341 | 147317235 | 1 | 15 | A | 0.0193 | 1.47 | 0.0577 | 1.46 | 0.1707 | 1.49 |
| rs74123747 | 147382534 | 1 | 16 | T | 0.0496 | 1.27 | 0.2194 | 1.20 | 0.0813 | 1.50 |

PPMS, primary progressive MS; BOMS, bout-onset MS (RRMS + SPMS); OR, odds ratio; LD#, linkage disequilibrium block, SWE, Swedish cohort; ITA, Italian cohort; CHR, Chromosome; P, P-Value; A1, effect allele; BP, base pair (GRCh37/hg19).
<sup>a</sup>SNP ID highlighted in bold remain significant after Bonferroni correction for multiple testing ( $p < 7.2 \times 10^{-4}$ ).

We next assessed whether the identified genetic variations that associate with PPMS and methylation impact gene expression in the brain tissue. Taking advantage of the GTEx and xQTL serve databases, we found that a large fraction of the variants (mapping to 10/16 and 34/57 LD blocks from the meta-analysis and Swedish cohort, respectively) conditioned transcript levels of neighboring genes within the locus *(PRKAB2, CHD1L* and *FMO5)*, as well as long non-coding RNAs in the brain (SI Appendix Table S7). As exemplified in Figure 3d, variants predisposing for higher DMR methylation and PPMS risk associated with lower *CHD1L* and *PRKAB2* expression while variants conferring low DMR methylation and PPMS protection associated with higher *CHD1L* and *FMO5* expression and lower *PRKAB2* transcript levels. *CHD1L* gene displayed the most consistent changes regarding the impact of genetic variation on PPMS risk, methylation and *CHD1L* expression. These data connecting the genetic variation, methylation and transcription in the extended 1q21.1 region are consistent with genomic annotations of the DMR underscoring putative regulatory features as well as short-range physical interaction with proximal chromatin segments harboring eQTL genes (SI Appendix Fig. S1). Thus, the variants influencing the risk of developing PPMS and controlling DNA methylation in the 1q21.1 locus likely also exert an effect on the expression of proximal genes in the brain, particularly *CHD1L, FMO5* and *PRKAB2*.

### Methylation at the 1q21.1 locus regulates proximal gene expression in neuron-like cells

In order to functionally address the causal relationship between epigenetic and transcriptional changes, we first tested whether methylation at the 1q21.1 DMR can exhibit regulatory properties on transcription, as suggested by genomic annotations of the locus (Fig. 2a, SI Appendix Fig. S1), by examining methylation-sensitive enhancer and promoter activity using in-vitro methylation assays. Two distinct 1 kb fragments harboring the DMR sequence, isolated from PPMS patients with low (rs1969869: CC) and high (rs1969869: AA) methylation levels at the identified DMR, were inserted in CpG-free vector-based reporter system. The sequences were subsequently methylated using two different methyltransferases, *M*.*SssI* enzyme methylating all CpG sites (57 CpGs) and *HhaI* enzyme targeting only internal cytosine residues from the consensus sequence GCGC (7 CpGs). The 3’-5’ oriented DNA sequence of both genotypes exhibited potent constitutive promoter activity on the reporter gene (Mock condition), this basal activity being nonetheless halved in carriers of the minor allele compared to AA-homozygotes (Fig. 4a). The DMR manifested enhancer activity as well, although to a lesser extent, with no clear contribution of each allele or sequence orientation (Fig. 4b). Comparison of the unmethylated and methylated sequences revealed a significant reduction of the promoter and enhancer activity when the insert was fully methylated (Fig. 4a-b). Thus, controlling the level of methylation in the DMR region might have the potential regulatory effect on gene expression.

**Figure 4.**
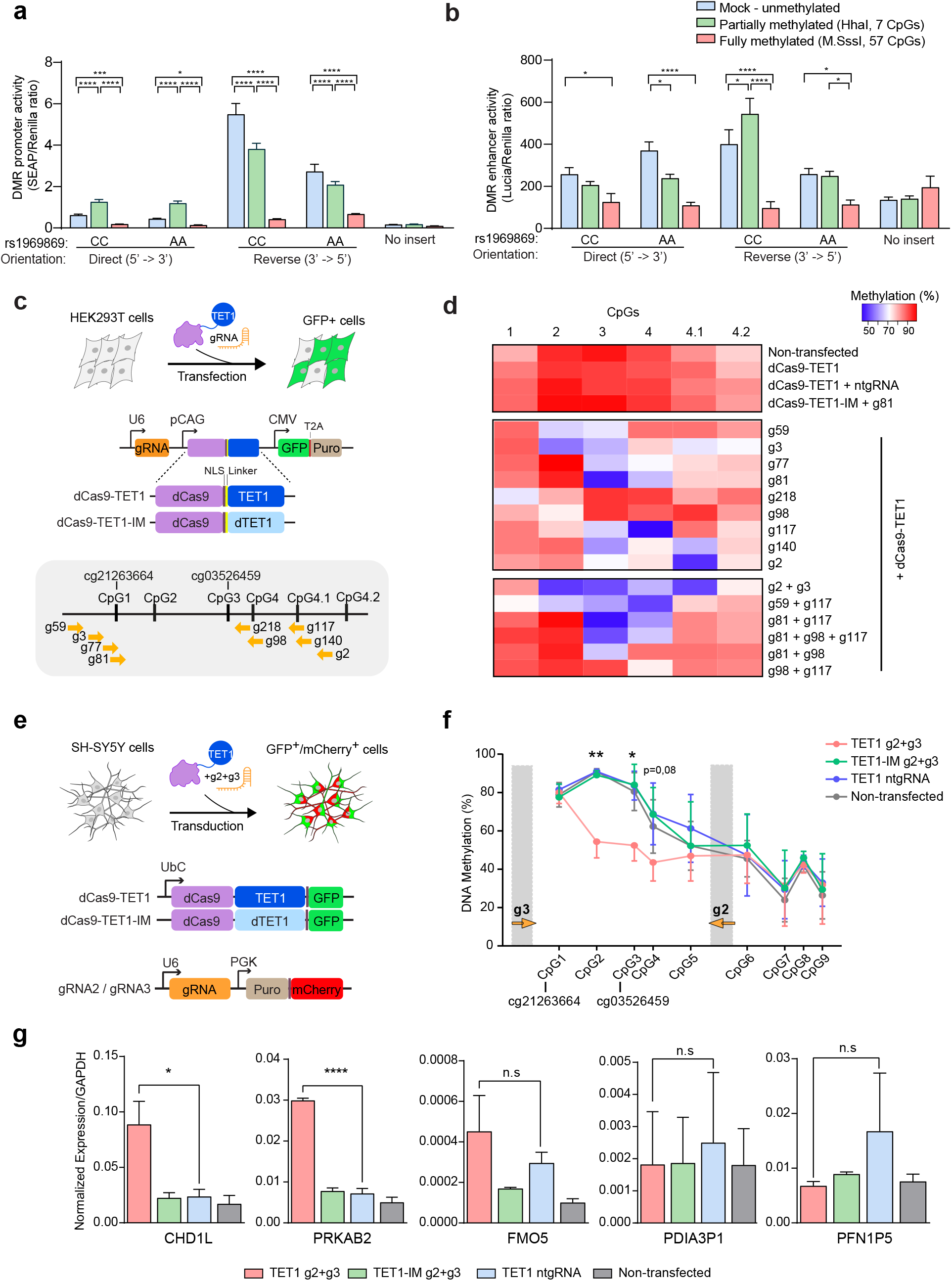
Functional impact of methylation at the 1q21.1 DMR on gene expression. **a-b**. Promoter (**a**) and enhancer (**b**) activity of the DMR, using CpG-free promoter-free (SEAP) and promoter-containing (Lucia) reporter gene vectors, respectively. Constructs in direct or inverted orientation of DNA segments derived from individual varying according to the genotype at rs1969869 were partially or fully methylated using *HhaI* and *M*.*SssI* enzymatic treatment, respectively. Results show relative activity (Lucia or SEAP normalized against Renilla) using three replicates in a representative experiment performed at least two times (2-way ANOVA test with Bonferroni correction for multiple comparisons (± SD)). **c**. Schematic representation of the experimental design for gRNAs screen in HEK293T cells including the features of the constructs and gRNA locations. **d**. Heatmap of the DNA methylation levels in successfully transfected (GFP positive) HEK293T cells three days following co-transfection of dCas9-TET1 with single or combined gRNAs in comparison to control conditions, deactivated TET1 (TET1-IM), non-targeting gRNA (ntgRNA) and non-transfected cells. **e**. Schematic representation of the experimental design for functional investigation in SH-SY5Y cells. **f**. DNA methylation levels in SH-SY5Y cells following delivery of dCas9-TET1-, gRNA2- and gRNA3-containing lentiviruses. Methylation percentages represent the average methylation levels of three experiments. **g**. Experiment showing expression of *CHD1L, PRKAB2, PDIA3P1* and *FMO5* genes relative to *GAPDH* transcript levels, quantified using RT-qPCR. The expression levels represent the average of three experiments (Two-tailed Student’s t test (± SD)). ****=p<0,0001, n.s = non-significant.

To formally address the impact of DNA methylation on endogenous gene expression, we exploited the CRISPR/dCas9-based epigenome editing platform which allows editing of methylation in a sequence-specific manner in combination with tailored single guide RNA (gRNA). We took advantage of the fact that several cell lines display hypermethylation at the 1q21.1 locus (as annotated by ENCODE Roadmap), mimicking methylation pattern in PPMS patients, and developed constructs expressing deactivated Cas9 fused to the catalytic domain of the demethylating enzyme TET1 (dCas9-TET1) to remove methylation from specific CpGs (see Methods for further description). We could validate the editing efficiency of dCas9-TET1 constructs by targeting 5 CpGs composing *MGAT3* gene promoter, with 10-50% de-methylation as previously reported (21) (SI Appendix Fig. S2). To remove methylation at CpGs of the PPMS-associated DMR, we first designed and functionally screened 9 gRNAs targeting the two 450K probes exhibiting the largest changes (cg21263664, cg03526459) and adjacent CpGs in HEK293T cells (Fig. 4c). Negative controls consisted of cells transfected with a non-targeting (nt) gRNA, the catalytically inactive form of TET1 (TET1-IM) or non-transfected cells. DNA methylation analysis, using pyrosequencing, showed different degrees of de-methylation ranging from 10-50% reduction in methylation at single or multiple CpG sites depending on the gRNA (Fig. 4d). Delivery of dCas9-TET1 together with gRNA2 and gRNA3 could achieve robust demethylation at four sites, including cg03526459, in HEK293T cells (Fig. 4d). We next adapted this strategy to neuroblastoma SH-SY5Y cells using transduction with dCas9-TET1 + gRNA2 + gRNA3 lentiviruses (Fig. 4e) and could confirm the editing efficiency in these cells with 15-35% decrease of DNA methylation spanning throughout CpGs 1-3 (Fig. 4f). We then assessed by qPCR the expression of genes located in the vicinity of the DMR. These include the three coding genes found regulated by the same meQTL-SNPs in the normal brain (Fig. 3c), namely *CHD1L* gene encoding a DNA helicase protein involved in DNA repair, *PRKAB2* gene encoding the non-catalytic subunit of the AMP-activated protein kinase (AMPK), which serves as an energy sensor protein kinase regulating cellular energy metabolism and *FMO5* gene encoding a Baeyer-Villiger monooxygenase implicated in liver function and metabolic ageing (22). The induced de-methylation, even moderate, of CpGs in the 1q21.1 DMR in SH-SY5Y cells resulted in increased expression of *CHD1L* and *PRKAB2* genes while the expression of *FMO5* and other proximal transcripts, *PDIA3P1* and *PFN1P5*, did not vary significantly (Fig. 4g). Of note, we did not observe any significant gene expression changes following dCas9-TET1 co-delivery with single or combined gRNAs in HEK293T cells (SI Appendix Fig. S3).

Collectively, these data suggest that the intergenic PPMS-associated DNA methylation at 1q21.1 region exerts regulatory properties on transcriptional processes. The methylation-mediated regulation of gene expression affects *CHD1L* and *PRKAB2* genes in neuron-like cells.

### Network analysis implicates *CHD1L* and *PRKAB2* genes in PPMS brain pathology

To further delineate the putative relevance of the genes included in the 1q21.1 locus in PPMS brain pathology, we constructed an unbiased correlation network analysis using RNA-sequencing-derived gene expression matrix from brain tissue samples of progressive MS patients (n = 12) and non-neurological controls (n = 10) (9) (Fig. 5). After gene-pair permutation and planar filtering, the network consisted of 0.5 million interactions among 27,059 genes (FDR < 0.05) (Fig. 5a). Multiscale clustering analysis further clustered these interactions into 757 non-overlapping modules (Fig. 5b). We then evaluated the relevance of these modules for PPMS (n = 5) and SPMS (n = 7) phenotypic traits by applying cluster trait association analysis (Fig. 5b). Three modules survived statistical testing, one of them (module #1) being correlated to SPMS trait while the two others (modules #2 and #3) showing high significance to PPMS phenotype (Fig. 5b, SI Appendix Table S9). Strikingly, these modules were found centered on genes from the extended 1q21.1 locus. Indeed, exploration of the biggest module (module #2), consisting of 380 nodes and 1079 interactions, revealed *CHD1L* as a central node within this network, closely interacting with six genes of a sub-module (Fig. 5c). Relative centrality could also be observed for the other two modules, which include *FMO5* among 40 nodes (module #1) and *PRKAB2* among 54 nodes (module #3) (Fig. 5c). Of note, *CHD1L* and *PRKAB2* were the only genes from the extended 1q21.1 locus to partake in the regulatory gene network underlying PPMS brain pathology in our analysis. Gene ontology analysis, using Ingenuity Pathway Analysis, underscored implication of all modules in nervous processes. This is particularly the case for module #2 with terms linked to neuronal functions, while additional *Biological Functions* were found associated to module #1 (e.g. metabolic processes) and module #3 (e.g. cell cycle) (Fig. 5c, SI Appendix Table S10). Thus, findings from unbiased *in*-*silico* approach support a potential role of genotype-methylation-expression regulatory network affecting genes of chromosome 1q21.1 extended locus, such as *CHD1L* and *PRKAB2*, in CNS processes in PPMS patients.

**Figure 5.**
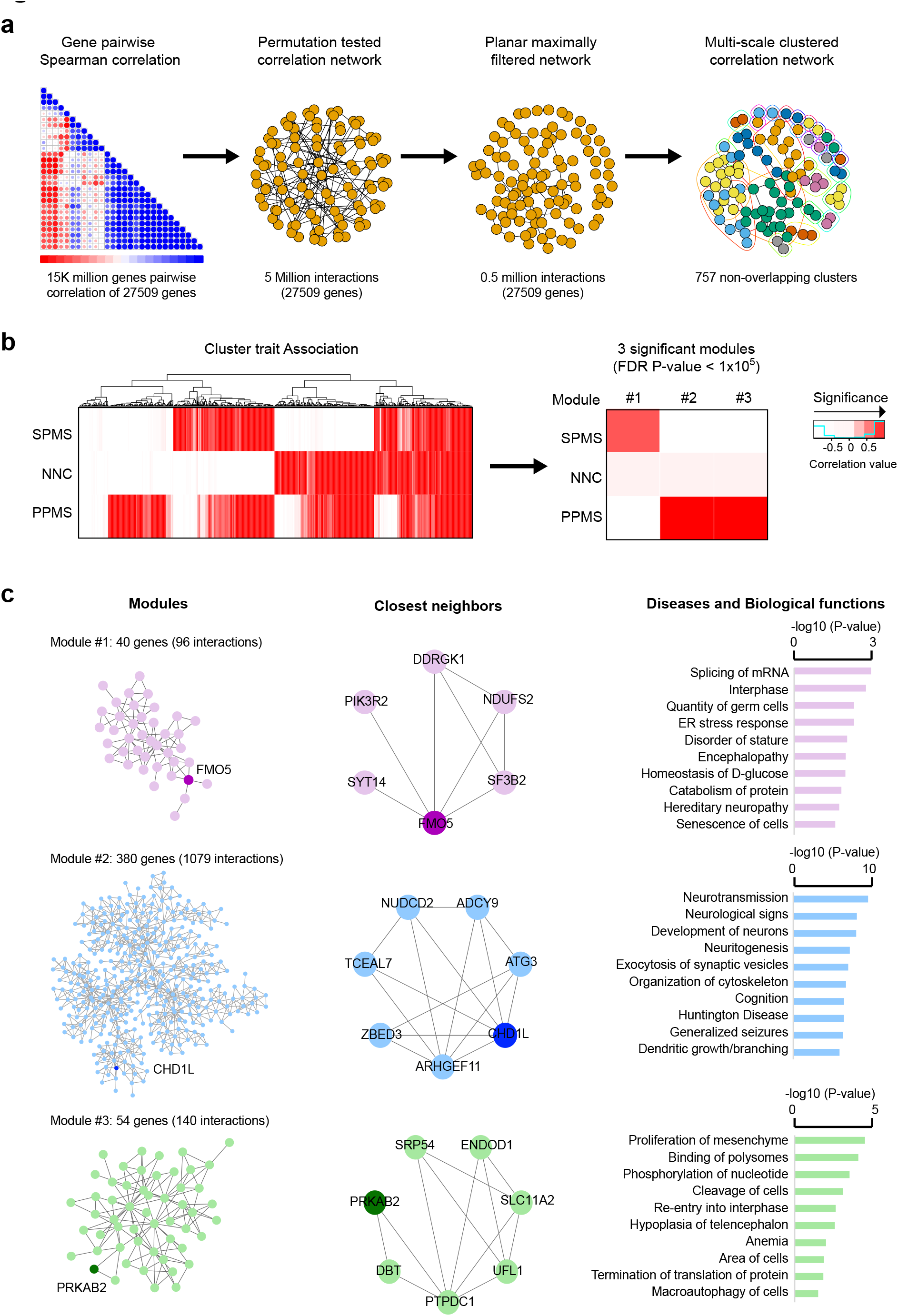
Implication of *CHD1L* and *PRKAB2* genes in PPMS brain pathology. **a**. Schematic representation of the correlation network analysis using bulk MS brain transcriptome(9). The different steps are depicted from left to right: correlation matrix containing gene-gene pairwise Spearman correlations; illustration of the permutation test filtered correlation network surviving an FDR P-value < 0.05 and the planar maximally filtering to convert the scale free network to be able to overlay on a plane spherical surface, reducing the network to 0.5 million interactions among 27509 genes and, finally; representation of the multiscale clustering of the network resulting into 757 non-overlapping clusters. **b**. Heatmap of the correlation coefficients of all 757 non-overlapping modules (left) and the three modules surviving correlation FDR P-value < 0.05 (right) to each tested phenotypic trait, with red gradient colors representing negative (not significant) to positive (significant) correlation. **c**. Representation of the three modules (left), the closest neighbors to candidate genes in each module (middle) and Gene Ontology findings, i.e. *Disease and Biological functions*, obtained using Ingenuity Pathway Analysis. MS, multiple sclerosis; PPMS, primary progressive MS; SPMS, secondary progressive MS; NNC, non-neurological controls.

To verify the network modules, we used large GTEx dataset from healthy individuals (SI Appendix Table S11). We did not detect our networks in any of 14 strata, but *CHD1L, FMO5* and *PRKAB2* genes belonged to significantly co-expressed genes networks only in the cortex and tibial nerve. This could indicate that, beyond constitutive expression of these genes in multiple tissues and cells, the existence of the networks is dependent on the disease processes and possibly more restricted to neurons. We thus investigated additional datasets comprising CNS tissue from MS patients, particularly focusing on those in which co-expressed networks containing *CHD1L, FMO5* or *PRKAB2* gene were detectable (SI Appendix Table S11).The CUX2^+^ excitatory neurons, previously shown to be specifically affected in MS, displayed significant co-expression of *CHD1L, FMO5* and *PRKAB2* with other genes, thereby forming network modules, in the single nuclei RNA sequencing (snRNA-seq) dataset from the original study (23). We detected a significant overlap of our *CHD1L* module, but not *FMO5* and *PRKAB2* modules, with the CUX2^+^ specific gene signature (overlapping = 18, p-value = 2.95 × 10^−6^, OR = 3.65). Conversely, the *CHD1L* module in CUX2^+^ neurons significantly overlapped with our *CHD1L* module with shared closest interacting genes and pathways related to neuronal functions (overlapping = 20, p-value = 0.03, OR=1.56, SI Appendix Table S9, SI Appendix Fig. S4).

## Discussion

MS is a multifactorial and heterogeneous neurological disease with unpredictable trajectories affecting young individuals. Treatment of progressive forms remains the greatest challenge in the care of MS patients, which is likely due to our partial knowledge of the pathological processes underlying these forms of disease. In this study, we utilized a multi-omics (genetic, epigenetic, transcriptomic) approach in several cohorts, in combination with in-vitro functional experiments and network analysis, to gain more insights into the primary progressive form of MS. We identified a genetically controlled hypermethylated region in the chromosome 1q21.1 locus which might affect the expression of proximal genes, particularly *CHD1L*, in the brain of PPMS patients.

Attempts to identify genetic variants that predispose for PPMS have not yielded reproducible associations with the exception of the known RRMS-risk *HLA-DRB1*15:01* variant (24, 25). One of the challenges of the genetic studies in PPMS is the limited availability of PPMS cases that comprise around 10% of all MS cases. We and others have shown that integrating additional layers of gene regulation, such as DNA methylation, can reveal genetic associations that are difficult to identify using conventional genetic studies (11, 26-28). In this study, we combined methylome profiling with locus-specific genetic association analyses in independent cohorts and identified a genetically controlled DMR in the 1q21.1 locus that displayed hypermethylation in PPMS patients specifically. Since SNPs can affect both CpGs as well as the probe/primer hybridization in methylation assays, we have ensured that DNA methylation measurements are not subject to such technical biases.

The same genetic variants that control methylation and gene expression in the locus associated with PPMS risk, suggesting a functional link between genetic, epigenetic, and transcriptional modalities in 1q21.1 in PPMS patients. We provided evidence for association with the risk of developing PPMS in two independent cohorts from Sweden and Italy comprising 1,104 PPMS cases compared to 11,335 of relapse onset MS, which is the largest reported PPMS cohort to date. Nevertheless, establishing unequivocal genetic association alone would require cohort sizes that could be challenging to obtain even by large multicentric efforts, further emphasising the need to combine multiple independent layers of evidence.

Our findings should also be considered in the light of the remarkably complex genomic architecture of the 1q21.1 locus displaying considerable copy-number variations. Indeed, the extended 1q21.1 genomic locus covering the identified DMR is characterized by the largest copy number expansion between non-human and human primate lineage, such variation correlating with evolutionary brain size in a dose-dependent manner (16, 17). Copy-number variations in this region have also been associated with brain disorders such cognitive and motor deficits, neuroblastoma, autistic spectrum disorder and schizophrenia (18-20). Our further analysis revealed that some of the SNPs conferring risk for PPMS, are associated with Schizophrenia (29) and are controlling the expression of neighboring genes including *CHD1L* in the brain cells. These associations with brain size and neurodevelopmental disorders further support the idea that inherent brain tissue vulnerability influenced by the genes in the locus may predispose for rapid and progressive MS disease, as recently suggested for the severity of MS (30). Whether the changes identified in the 1q21.1 locus in our study are related to structural variations of the genome warrants further investigation.

The observed dual meQTL and eQTL effects in the CNS tissue, underscore the possible contribution of genetic-epigenetic-transcriptional regulation in PPMS brain pathology. In this context, we found that genetic variation that strongly associated with elevated DNA methylation levels at 1q21.1 and modestly associated with increased PPMS risk is linked to lower expression of *CHD1L* transcript in the CNS. We next sought to formally explore the possibility that DNA methylation may directly influence gene expression. We first demonstrated intrinsic methylation-sensitive promoter and enhancer activity of PPMS-associated DMR sequence using in-vitro methylation assays. We then utilized CRISPR/dCas9-TET1 epigenome editing approach to achieve effective and locus-specific demethylation at CpGs of the identified DMR and found that manipulating DNA methylation at the 1q21.1 DMR affects the expression of the proximal *CHD1L* and *PRKAB2* genes in neuron-like cells, an effect we could not observe in HEK293T cells. These data, together with the indisputable involvement of 1q21.1 region in neuronal circuitry and brain integrity (16-20), implicate genes of the locus in neuropathological processes in PPMS. Undeniably, given the occurrence of meQTL effects in the blood, one cannot exclude a contribution of 1q21.1 regulatory network in the peripheral immune compartment as well. Similar trans-tissue effects with shared pattern between the immune and nervous compartments have previously observed in the context of progressive MS disease (31). Collectively, these findings portray DNA methylation as a putative intermediary of genetic influence on gene expression, particularly in the CNS of PPMS patients.

Our unbiased correlation network analysis of published transcriptomic data obtained from post-mortem brain tissue from PPMS, SPMS patients and controls (9), identified *CHD1L* and *PRKAB2* transcripts as implicated in PPMS specific regulatory gene networks while *FMO5* gene was found significantly connected to SPMS phenotype. *FMO5* encodes for an atypical flavin containing monooxygenase protein with the ability to oxidize a wide range of substrates and xenobiotic, whose disruption has been shown to affect metabolic ageing in rodents (32). *CHD1L* encodes a DNA helicase implicated in chromatin-remodelling during DNA repair, with both depletion and overexpression of CHD1L leading to genomic sensitivity to DNA-damaging agents (33). *PRKAB2* encodes the beta subunit of AMP-activated kinase, a neuroprotective energy-sensing kinase exerting pivotal roles in the cellular energy metabolism required for maintenance of neuronal integrity (34). Accordingly, previous studies on neuron-specific loss of PRKAB2 function fruit flies resulted in progressive fragmentation of dendritic arborisation, cognitive impairment as well as reduced stress resistance and lifespan (34). Interestingly, bulk and single cell transcriptomics in brain tissue have found dysregulation of several genes located within the identified 1q21.1 locus, including *CHD1L, PRKAB2, FMO5* transcripts along with several members of the *NBPF* family of genes, in progressive MS patients compared to control individuals (9, 23). Markedly, *CHD1L* gene has been identified in the stress-related signature of excitatory CUX2-expressing projection neurons in upper-cortical layers, which are particularly susceptible to degeneration in MS patients (23). Additionally, a large fraction of *CHD1L* (3/6) closest network neighbours identified in our study were found significantly upregulated in these stressed excitatory neurons as well. Moreover, our *CHD1L* network module displayed a significant overlap, both on the gene and pathway level, with the transcriptional signature of CUX2^+^ neurons as well as the *CHD1L* network detected in CUX2^+^ neurons using snRNA-seq dataset from Schirmer et *al*. (23). It is therefore reasonable to speculate on the involvement of *CHD1L* in the compensatory mechanisms counteracting the cellular burden, e.g. genomic and metabolic stress, observed in these neurons, although validation in large PPMS dataset is warranted. More specifically, genetic predisposition to high methylation and low expression of *CHD1L* gene might influence susceptibility to PPMS by limiting the inherent repair capacity in the brain.

In conclusion, our study suggests that PPMS patients display distinct molecular changes compared to SPMS, RRMS and healthy individuals. The data further support the hypothesis of a causal genetic-epigenetic-transcriptional interplay within the extended 1q21.1 locus, with functional evidence of contribution from *CHD1L* in progressive MS pathology. Given the reversible nature of DNA methylation, our findings open new avenues for development of therapeutic strategies, such as the targeted epigenetic therapy, in the treatment of progressive MS.

## Materials and Methods

### Cohorts

Details of the cohorts are described in the SI Appendix Table S1. Briefly, for genome-wide DNA methylation and methylation quantitative trait locus (meQTL) analysis used in cohort 1, peripheral blood samples were collected form 140 MS, patients including 120 RRMS, 4 PPMS and 16 SPMS patients, and 139 healthy individuals (HC), as previously described (11). An independent cohort, cohort 2, consisting of 48 RRMS and 36 PPMS patients (matched for age, sex, disease duration and Swedish descent) was used for pyrosequencing validation and locus-specific meQTL analysis (35). Cohort 3 and the additional Italian cohort used for genetic association study comprises 1,104 PPMS and 11,335 BOMS, as described in the corresponding sections below. Gene expression data (RNA-sequencing) from bulk brain tissue samples of progressive MS patients (5 PPMS and 7 SPMS) and non-neurological controls (n = 10), previously described (9), were used for correlation network analysis and validation was performed in neuronal snRNA-seq data (23). Additionally, we utilized publicly available databases from xQTL serve (15) (n = 543 bulk prefrontal cortex samples, http://mostafavilab.stat.ubc.ca/xQTLServe/) and GTEx (selecting all available nervous tissues, https://gtexportal.org/) platforms to address meQTL and eQTL effects in the CNS. No sample size calculation was performed for the cohorts involved in this study.

### Ethics approval and consent to participate

All experiments on human subjects were approved by the Regional Ethical Review Board in Stockholm and carried out in accordance with institutional guidelines. Written informed consent was obtained from all study participants.

### Genome-wide DNA methylation and meQTL analyses (cohort 1)

#### DNA methylation analysis

The methylation data from Infinium HumanMethylation450 (450K) arrays was preprocessed as previously described (11) and in supporting information using the Illumina default procedure implemented in the Bioconductor minfi package (36) To identify differentially methylated regions (DMRs) associated with the PPMS phenotype, we used the bumphunter function in minfi package (36) with adjustment for confounders: age, sex, self-reported smoking status (ever smokers vs. never smokers), hybridization date, and the first two principle components of estimated differential cell counts. Region that has a family wise error rate (FWER) less than 0.05 with 1000 resamples and contains at least 2 probes was identified as a trait-associated DMR.

#### Methylation QTL analysis

To identify potential genetic dependency, the PPMS-associated DMR was tested for association with genotype (594,262 SNPs) using an additive minor-allele dosage model. Genotype-DMR associations were corrected for multiple testing using a stringent Bonferroni-adjusted threshold of 0.05.

### Locus-specific DNA methylation and meQTL analyses (cohort 2)

#### DNA methylation analysis

For validation of the identified PPMS-associated DMR, pyrosequencing analysis of 7 CpG sites in the locus was performed as detailed in supporting information and SI Appendix Table S12, SI Appendix Figure S5.

#### Methylation QTL analysis

Methylation data was RANK transformed in R using R Core team (Vienna, Austria, https://www.R-project.org/). Genotyping was carried out at deCODE (deCODE genetics/Amgen, Reykjavik, Iceland) using Illumina OmniExpress chip with 716,503 SNPs mapped to the Human Assembly Feb.2009 (GRCh37/hg19). Of 84 individuals, 83 where genotyped in deCODE and 82 of them passed QC. We performed meQTL analysis of chromosome 1 from bp 146500000 to bp 147000000, in PLINK (37) excluding SNPs with less than 98% genotyping rate and SNPs that were not in Hardy-Weinberg equilibrium (p < 0.05) and corrected for 5 population based (ancestral informative markers) principal component analysis covariates. After quality control, 123 SNPs remained in the region. Genotype-CpG associations were corrected for multiple testing using stringent Bonferroni-adjusted threshold of 0.05.

### Genetic association study in the Swedish (SWE) cohort

Patients from the Swedish (SWE) cohort were genotyped in two different batches at deCODE Genetics using Illumina Human OmniExpress 24 v1 (OE) and Global Screening Array MD 24 v2 (GSA) arrays, following manufacturer’s instructions. For the genetic association analysis we extracted all the imputed SNPs (n = 3,057) in the extended chr1 locus (from bp 146500000 to bp 147000000). A total of 7,682,164 autosomal variants passed quality controls and were tested for SNP-to-phenotype association (PPMS = 603 versus BOMS = 9,247) in the generalized linear model analysis including covariates, separately for individuals genotyped on OE chip and those genotyped on GSA arrays (see details in supporting information). Subsequently, the results from the two independent association studies (OE and GSA) were meta-analyzed using fixed-effect and random-effect models as implemented in PLINK. Sixty-nine LD blocks were identified and used in a Bonferroni correction to account for multiple testing.

### Genetic association study in the Italian (ITA) cohort

For the Italian cohort, patients were recruited at the Laboratory of Human Genetics of Neurological Disorders at the San Raffaele Scientific Institute in Milan, Italy and genotyped on Illumina platforms. After quality controls (see supporting information), a logistic regression model, as described for the SWE cohort, was used to study the association between the SNPs in the extended chr1 locus and the course of MS in a total of 2,589 patients (PP = 501; BOMS = 2,088). Sex and PC 1 to 8 were used as covariates in the model.

### SWE and ITA meta-analysis

A fixed-effect model meta-analysis of the standard errors of the odds ratio, as implemented in Plink (38), was applied on the three cohorts. The number of common variants in all the cohorts was 2,676. Multiple testing issue was addressed as described for the SWE cohort (see also supporting information).

### In-vitro methylation assay

To address the regulatory features of the identified DMR, we used in-vitro DNA methylation reporter assay. A 927 bp fragment encompassing the identified DMR was amplified using blood genomic DNA from PPMS patients presenting with low (rs1969869: CC) and high (rs1969869: AA) methylation levels at the identified DMR. The amplified products in direct and reverse orientation were inserted into pCpG-free promoter vector (Invivogen) containing a Lucia luciferase reporter and into a pCpG-free basic vector (Invivogen) containing a murine secreted embryonic alkaline phosphatase (mSEAP) reporter gene for assessment of enhancer and promoter activity, respectively. After complete, partial or mock methylation of the constructs and transfection of HEK293T cells, Lucia or SEAP signals were normalized against Renilla (triplicate) and experiments were replicated at least two times. Details are provided in supporting information.

### CRISPR/dCas9-TET1 epigenome editing

#### dCas9-TET1 and gRNA generation

To address dCas9-TET1-mediated epigenome editing in SH-SY5Y cell line, we utilized the lentivirus version of the cassettes. Detailed methods to engineer the P3-dCas9-Tet1-GFP-Puro (Addgene #190728), P3-Lenti-dCas9-Tet1-GFP (Addgene #190729) and P3-Lenti-dCas9-Tet1IM-GFP (Addgene #190730) constructs are available in supporting information. Details and maps of the final constructs used in this study are presented in SI Appendix Figure S7. All gRNAs were designed by CRISPOR Version 4.98 (39) both on the sense and antisense strands, with sequence and mapping presented in the SI Appendix Table S12 and SI Appendix Fig. S8.

#### dCas9-TET1 delivery to HEK293T and SH-SY5Y cells

To test the efficiency of epigenome editing, we exploited HEK293T ease of transfection and performed gRNAs screen. Different gRNAs were transfected either individually or in combination based on the target site, using Lipofectamine 3000 (Invitrogen). For all experiments both on HEK293T and SH-SY5Y cells, DNA was extracted after 72 hours and bisulfite conversion was performed using 200 ng of the extracted DNA (BS-DNA, EZ DNA methylation kit, ZYMO research). DNA methylation was assessed using pyrosequencing, as described above. For all experiments conducted on SH-SY5Y cells, we delivered a mix of the two gRNAs that showed the highest efficiency in reducing methylation in HEK293T cells (gRNA #2 and #3). Lentiviruses generation and subsequent transduction of SH-SY5Y cells are performed based on the standard protocols which details are available in supporting information.

### qPCR analysis

Total RNA and DNA were extracted using AllPrep DNA/RNA Kits (Qiagen) according to the manufacture instruction. RNA and DNA concentrations and quality were verified by QIAxpert (Qiagen). Reverse transcription of RNA was performed using the manufacturer’s instructions of iScript™ cDNA Synthesis Kit (Bio-Rad Laboratories, Inc., CA). Real-time PCR was performed on a BioRad CFX384 Touch Real-Time PCR Detection System using iQ™ SYBR® Green Supermix (Bio-Rad Laboratories, Inc., CA) in a standard three step PCR protocol. The relative expressions of the selected genes were normalized to the reference gene *GAPDH*. The specificity of real time PCR reaction was verified by the melt curve analysis. The expression level of selected genes were analyzed using ΔΔCT method (40) and compared via independent t-test. All statistical analyses were performed in GraphPad Prism 6 and 7 (GraphPad Software).

### Correlation network analysis in MS brain

#### Raw data analysis

The fastq files corresponding to bulk gene expression (RNA-sequencing) data from brain tissue samples of progressive MS patients (n = 12) and non-neurological controls (n = 10) (9, 23) were extracted from the RAW RNA sequence files and checked for quality control using multiqc software to make them ready for alignment (41). After trimming using the trimgalore program (42), fastqc files were aligned and annotated using STAR aligner and Stringtie software (43) by applying human hg38 refseq information from UCSC. The analysis was performed on the extracted count matrix using bash and Python.

#### Network analysis

In order to utilize a brain-specific network module, we applied a previously established bioinformatic pipeline utilizing co-expression network analysis (44), as described in supporting information. The final network consisted of 0.5 million interactions among 27,059 expressed genes which clustered into 757 non-overlapping modules. For the validation data, we applied the same pipeline on several datasets (SI Appendix Table S11). In the CUX2^+^ neuronal snRNA-seq count data (23), planar maximal filtration of the Spearmen correlated network of 10780 genes was multiscale clustered using the MEGENA package in R. This resulted in 91 modules out of which 1 module with *CHD1L* was significant and was further analyzed with Fisher enrichment test and pathway analysis using clusterProfiler.

#### Cluster Trait association analysis

Principal component analysis (PCA) is first performed for each cluster. Next, correlation between the first principal component and each trait was computed as cluster relevance to the trait. The 757 clusters identified from the correlation network were evaluated for the relevance to PPMS, SPMS and control phenotypes. Three clustered passed FDR P-value < 0.05.

## Data Availability

The 450K array methylome data generated from whole blood (cohort 1) are available in the Gene Expression Omnibus (GEO) database under accession number GSE106648. The RNA-sequencing data used for correlation network analysis in MS brain are accessible under the accession numbers GSE174647, GSE118257, GSE179427, PRJNA544731 with details presented in Supplementary Table S11. Results from the genetic association analysis (cohort 3) are available in SI Appendix Tables. Due to the GDPR regulations, we cannot deposit any personal information including genetic data, which can be shared upon request and under a Data Access Agreement.

https://www.ncbi.nlm.nih.gov/geo/query/acc.cgi?acc=GSE106648

## Acknowledgments and funding sources

This study was supported by grants from the Swedish Research Council, the Swedish Association for Persons with Neurological Disabilities, the Swedish Brain Foundation, the Swedish MS Foundation, the Stockholm County Council - ALF project, the European Union’s Horizon 2020 research, innovation programme (grant agreement No 733161) and the European Research Council (ERC, grant agreement No 818170), the Knut and Alice Wallenberg Foundation, Hedlund Foundation, Åke Wiberg Foundation and Karolinska Institute’s funds. LK is supported by a fellowship from the Margaretha af Ugglas Foundation. MPK is supported by McDonald Fellowship from Multiple Sclerosis International Federation (MSIF) and the EU/EFPIA/OICR/McGill/KTH/Diamond Innovative Medicines Initiative 2 Joint Undertaking (EUbOPEN grant agreement No 875510). CSC is supported by the Blanceflor Boncompagni Ludovisi, née Bildt Foundation. PC is supported by NIH R35-NS111604. The funders of the study had no role in study design, sample acquisition, data collection, data analysis, data interpretation or writing of the manuscript. We are grateful to all volunteers for contributing to the study. We thank Maria Kalomoiri, Jessica Aguilar and Jacqueline Hammer for their help in experimental setup, Dr. Ewoud Ewing for statistics assistance, Dr. Alexander Espinosa for providing plasmid construct and Prof. Lucas Schirmer for providing the CUX2^+^ specific data. We acknowledge KIGene facility at the Center for Molecular Medicine and Karolinska Institutet for Sanger sequencing service. The computations were enabled by resources provided by the Swedish National Infrastructure for Computing (SNIC) through Uppsala Multidisciplinary Center for Advanced Computational Science (UPPMAX), partially funded by the Swedish Research Council through grant agreement no. 2018-05973. We thank deCODE for genotyping of the Swedish population (https://www.decode.com/).

