## Supplementary material for "Implication of DNA methylation changes at chromosome 1q21.1 in the brain pathology of Primary Progressive Multiple Sclerosis": SI_Appendix

**This PDF file includes:**

Detailed Materials and methods
Figures S1 to S8
SI References

### Detailed Materials and methods

### Materials and Methods

#### Cohorts

Details of the cohorts are described in the SI Appendix Table S1. Briefly, for genome-wide DNA methylation and methylation quantitative trait locus (meQTL) analysis used in cohort 1, peripheral blood samples were collected form 140 MS, patients including 120 RRMS, 4 PPMS and 16 SPMS patients, and 139 healthy individuals (HC), as previously described (1). An independent cohort, cohort 2, consisting of 48 RRMS and 36 PPMS patients (matched for age, sex, disease duration and Swedish descent) was used for pyrosequencing validation and locus-specific meQTL analysis (2). Cohort 3 and the additional Italian cohort used for genetic association study comprises 1,104 PPMS and 11,335 BOMS, as described in the corresponding sections below. Gene expression data (RNA-sequencing) from bulk brain tissue samples of progressive MS patients (5 PPMS and 7 SPMS) and non-neurological controls (n = 10), previously described (3), were used for correlation network analysis and validation was performed in neuronal snRNA-seq data (4). Additionally, we utilized publicly available databases from xQTL serve (5) (n = 543 bulk prefrontal cortex samples, http://mostafavilab.stat.ubc.ca/xQTLServe/) and GTEx (selecting all available nervous tissues, https://gtexportal.org/) platforms to address meQTL and eQTL effects in the CNS. No sample size calculation was performed for the cohorts involved in this study.

#### Ethics approval and consent to participate

All experiments on human subjects were approved by the Regional Ethical Review Board in Stockholm and carried out in accordance with institutional guidelines. Written informed consent was obtained from all study participants.

#### Genome-wide DNA methylation and meQTL analyses (cohort 1)

*DNA methylation analysis.* The methylation data from Infinium HumanMethylation450 (450K) arrays was preprocessed as previously described (1) using the Illumina default procedure implemented in the Bioconductor minfi package (6). Briefly, all samples were normalized together using the minfi preprocessQuantile function. The probe level raw data for each sample were normalized using Illumina's control probe scaling procedure and converted to methylation β values on the 0–1 scale (M/(M + U + 100), where M and U represent the methylated and unmethylated signal intensities, respectively). Cell counts for the six major cell types in blood (granulocytes, B cells, $CD4^+$ T cells, $CD8^+$ T cells, monocytes, and NK cells) for each individual were estimated using the estimateCellCounts function in minfi package (6), which obtain sample-specific estimates of cell proportions based on reference information on cell-specific methylation signatures (7). Results from the estimation can be found in (1). To identify differentially methylated regions (DMRs) associated with the PPMS phenotype, we used the bumphunter function in minfi package (6) with adjustment for confounders: age, sex, self-reported smoking status (ever smokers vs. never smokers), hybridization date, and the first two principle components of estimated differential cell counts. Region that has a family wise error rate (FWER) less than 0.05 with 1000 resamples and contains at least 2 probes was identified as a trait-associated DMR.

*Methylation QTL analysis.* To identify potential genetic dependency, the PPMS-associated DMR was tested for association with genotype (594,262 SNPs) using an additive minor-allele dosage model. Genotype-DMR associations were corrected for multiple testing using a stringent Bonferroni-adjusted threshold of 0.05.

### Locus-specific DNA methylation and meQTL analyses (cohort 2)

*DNA methylation analysis.* For validation of the identified PPMS-associated DMR, pyrosequencing analysis was performed using 500 ng of genomic DNA samples previously converted to bisulfite DNA (BS-DNA, EZ DNA methylation kit, ZYMO research) with PyroMark Q96 system (Qiagen). Primers and probes for three sequencing assays covering 7 CpG sites in the locus were designed by PyroMark Design software (Qiagen) (SI Appendix Table S12, SI Appendix Fig. S5). Around 10 ng of BS-DNA was amplified using PyroMark PCR kit (Qiagen) and the forward and 5′-biotinylated reverse primers. The entire PCR product, 4 pmol of the sequencing probe and streptavidin sepharose high-performance beads (GR Healthcare) were used for pyrosequencing on a PyroMark Q96 ID pyrosequencing instrument (Qiagen) using the PyroMark Gold 96 Reagent kit (Qiagen). Methylation levels were determined by the ratio of C and T by the PyroMark CpG software 1.0.11 (Qiagen) and expressed as percentage methylation at each CpG site. To verify the efficiency and sensitivity of the PCR-pyrosequencing, we used standard curves with unmethylated and methylated human BS-DNA samples (Qiagen). To test the differences in DNA methylation between PPMS and RRMS patients for each CpG site, non-parametric Mann-Whitney U test was applied with GraphPad Prism software (PRISM 7.0; GraphPAD Software Inc., San Diego, CA, USA). We have done extensive SNP and sequence analyses to assure that the methylation measurements of the majority of the DMR CpGs are not a result of the technical measurement artefacts driven by a potential effect of SNPs on the CpGs (8, 9), pyrosequencing or 450K assays.

*Methylation QTL analysis*. Methylation data was RANK transformed in R using R Core team (Vienna, Austria, https://www.R-project.org/). Genotyping was carried out at deCODE (deCODE genetics/Amgen, Reykjavik, Iceland) using Illumina OmniExpress chip with 716,503 SNPs mapped to the Human Assembly Feb.2009 (GRCh37/hg19). Of 84 individuals, 83 where genotyped in deCODE and 82 of them passed QC. We performed meQTL analysis of chromosome 1 from bp 146500000 to bp 147000000, in PLINK (10) excluding SNPs with less than 98% genotyping rate and SNPs that were not in Hardy-Weinberg equilibrium ($p < 0.05$) and corrected for 5 population based (ancestral informative markers) principal component analysis covariates. After quality control, 123 SNPs remained in the region. Genotype-CpG associations were corrected for multiple testing using stringent Bonferroni-adjusted threshold of 0.05.

### Genetic association study in the Swedish (SWE) cohort

Patients from the Swedish (SWE) cohort were genotyped in two different batches at deCODE Genetics using Illumina Human OmniExpress 24 v1 (OE) and Global Screening Array MD 24 v2 (GSA) arrays, following manufacturer's instructions. The cohort was aligned to the forward strand of the hg19 reference genome based on strand information from the Illumina array manifest files. Samples with less than 95% genotyping yield were excluded. Samples with a mismatch between reported and genetic sex were also excluded using a linkage disequilibrium (LD) pruned set of high-quality chromosome X variants. Genetic variants were filtered based on several criteria, including genotype missingness, Hardy-Weinberg Equilibrium (HWE), minor allele frequency (MAF), and differential missingness between cases and controls. Palindromic variants were also filtered based on alternate allele frequency. Individuals with an absolute inbreeding coefficient greater than 0.05 or relatedness at the third degree or closer were excluded. To control for population stratification prior to imputation, Principal Components (PC) were calculated, and outliers were excluded. The Haplotype Reference Consortium (HRC; version 1.1) imputation reference panel 8 was used for phasing and imputation of genotyping data (11). A total of 7,682,164 autosomal variants passed quality controls.

Starting from these, for the genetic association analysis we extracted all the imputed SNPs (n = 3,057) in the extended chr1 locus (from bp 146500000 to bp 147000000). A Principal Component Analysis (PCA) was performed using whole-genome autosomal markers after LD pruning (100 bp window size, 2 bp step, pairwise r2 threshold of 0.1). The first 8 PC and biological sex were included as covariates in the generalized linear model analysis of SNP-to-phenotype association (PPMS =

603 versus BOMS = 9,247, to account for residual population stratification. The PCA and the association analyses were run separately for individuals genotyped on OE chip and those genotyped on GSA arrays, to minimize the batch effect exerted by array architecture and potential differences in recruitment of individuals between the two datasets. Subsequently, the results from the two independent association studies were meta-analyzed using fixed-effect and random-effect models as implemented in PLINK. To gain an insight on the underlying genetic structure, we estimated the haplotype blocks in the extended chr1 locus using the largest cohort (OE cohort) as reference on SNPs with MAF => 0.05 adopting the standard method integrated in PLINK (12). Sixty-nine LD blocks were identified and used in a Bonferroni correction to account for multiple testing.

**Genetic association study in the Italian (ITA) cohort**

For the Italian cohort, patients were recruited at the Laboratory of Human Genetics of Neurological Disorders at the San Raffaele Scientific Institute in Milan, Italy and genotyped on Illumina platforms. Prior to imputation, we excluded subjects for which sex mismatch, those with call-rate < 90% and outliers exceeding the mean level of heterozygosity by > 3 standard deviations. At variant level, we discarded rare SNPs with MAF < 1%, SNPs with a call-rate < 90% and those departing from HWE at $p < 10 \times 10^{-6}$. Imputation was carried out to HRC reference genome (11). A logistic regression model, as described for the SWE cohort, was used to study the association between the SNPs in the extended chr1 locus and the course of MS in a total of 2,589 patients (PP = 501; BOMS = 2,088). Sex and PC 1 to 8 were used as covariates in the model.

**SWE and ITA meta-analysis**

A fixed-effect model meta-analysis of the standard errors of the odds ratio, as implemented in Plink (13), was applied on the three cohorts. The number of common variants in all the cohorts was 2,676. Multiple testing issue was addressed as described for the SWE cohort.

**In-vitro methylation assay**

To address the regulatory features of the identified DMR, we used in-vitro DNA methylation reporter assay. A 927 bp fragment encompassing the identified DMR was amplified using primers containing overhanging *SpeI* and *NsiI* restriction sites (SI Appendix Table S12). We used blood genomic DNA from PPMS patients presenting with low (rs1969869: CC) and high (rs1969869: AA) methylation levels at the identified DMR. The amplified products in direct and reverse orientation were inserted into pCpG-free promoter vector (Invivogen) containing a Lucia luciferase reporter and into a pCpG-free basic vector (Invivogen) containing a murine secreted embryonic alkaline phosphatase (mSEAP) reporter gene for assessment of enhancer and promoter activity, respectively. As the body of these vectors is devoid of any CpGs, any impact of DNA methylation on reporter gene expression is restricted to the inserted fragment only. All the constructs were either completely methylated (57 CpGs) using *M.SssI* or partially methylated (7 CpGs residing in the GCGC sequence) by *HhaI* methyltransferases (New England BioLabs) using 1 ug of the vectors and 1 unit of the enzymes. The mock methylated control was treated equally but in absence of any methyltransferases and corresponds to unmethylated inserts. After the purification of the methylated, partially- and mock-methylated constructs (QIAquick PCR purification Kit, Qiagen), the efficiency of methylation was assessed using an EpiJET DNA Methylation analysis Kit (*MspI*/*HpaII*) (ThermoFisher Scientific), followed by gel electrophoresis (SI Appendix Fig. S6). Original vectors treated by *M.SssI* and *HhaI* or mock-treated were used as controls. Human embryonic kidney HEK293T cells were cultured in Dulbecco's Modified Eagle's medium in 96 well plates and co-transfected with 90 ng of the Lucia or SEAP constructs and 5 ng of the control vector pGL4-TK-hH Luc constitutively expressing Renilla luciferase, using Lipofectamine 3000 Transfection Reagent (Qiagen). Approximately, 48 hours post transfection, Lucia, SEAP and Renilla activities were measured using QUANTI-Luc (Invivogen), the Phospha-Light System (Applied Biosystems) and the Dual-Glo Luciferase Assay System (Promega), respectively, according to manufacturer's instructions, on the GloMax 96 Microplate Luminometer (Promega). Both direct and reverse orientations of the sequence were tested. Lucia or SEAP signals were normalized against Renilla (triplicate) and experiments were replicated at least two times.

**CRISPR/dCas9-TET1 epigenome editing**

*dCas9-TET1 and gRNA generation*. Details and maps of the final constructs used in this study are presented in SI Appendix Figure S7. Briefly, we engineered a P3-dCas9-Tet1-GFP-Puro (Addgene #190728) construct as follows. First, to be able to express the gRNAs from the same vector, we mutated the *BbsI* sites in the TET1 sequence (without changing the protein sequence) synthesized by Eurofins (Eurofins MWG Operon Ebersberg, Germany). We then utilized the backbone of a pdCas9-DNMT3A-EGFP plasmid (Addgene #71666) (14) and replaced DNMT3A with TET1 sequence. This cassette was previously engineered to express the original EGFP sequence in a double reporter cassette containing EGFP-T2A-Puromycin under the control of an independent CMV promoter to allow sufficient expression of GFP signal for post-transfection cell sorting. We proceeded similarly with the TET1-IM construct which expresses a deactivated TET1 catalytic unit. The final plasmid expressed dCas9-TET1 (or dCas9-TET1-IM) and CMV-EGFP-T2A-Puromycin double marker unit. All gRNAs were designed by CRISPOR Version 4.98 (15) both on the sense and antisense strands, with sequence and mapping presented in the SI Appendix Table S12 and SI Appendix Fig. S8.

To address dCas9-TET1-mediated epigenome editing in SH-SY5Y cell line, we utilized the lentivirus version of the cassettes. We used Fuw-dCas9-Tet1CD (Addgene #84475) and Fuw-dCas9-Tet1CD_IM (Addgene #84479) plasmids and added the EGFP marker to facilitate sorting of positively transduced cells (new plasmids named as P3-Lenti-dCas9-Tet1-GFP (Addgene #190729) and P3-Lenti-dCas9-Tet1IM-GFP (Addgene #190730)). We used pKLV2-U6gRNA3(BbsI)-PGKpuro2ABFP (Addgene #67990) to express the gRNA and replaced the BFP with mCherry (named as P3-pKLV2-U6gRNA(BbsI)-PGKpuro2A-mCherry). All constructs were confirmed by Sanger sequencing (KiGene) and chromatograms were analyzed using SnapGene software 5.2.3 (GSLBiotech). Plasmids constructed for this study can be obtained from Addgene.

*dCas9-TET1 delivery to HEK293T and SH-SY5Y cells*. To test the efficiency of epigenome editing, we exploited HEK293T ease of transfection and performed gRNAs screen. Different gRNAs were transfected either individually or in combination based on the target site, using Lipofectamine 3000 (Invitrogen). For all experiments both on HEK293T and SH-SY5Y cells, DNA was extracted after 72 hours and bisulfite conversion was performed using 200 ng of the extracted DNA (BS-DNA, EZ DNA methylation kit, ZYMO research). DNA methylation was assessed using pyrosequencing, as described above. For all experiments conducted on SH-SY5Y cells, we delivered a mix of the two gRNAs that showed the highest efficiency in reducing methylation in HEK293T cells (gRNA #2 and #3).

For the generation of lentiviruses and subsequent transduction of SH-SY5Y cells, HEK293T cells were seeded (~40-50 % confluent) in 2 ml DMEM with 10% serum and 2 mM glutamine per well in 6-well plates. The next day, cells had reached 60-70 % confluency and were co-transfected with transfer plasmids as P3-Lenti-dCas9-Tet1-GFP (Addgene #190729) or P3-Lenti-dCas9-Tet1IM-GFP (Addgene #190730), as well as P3-pKLV2-U6gRNA(BbsI)-PGKpuro2A-mCherry, psPAX2 (Addgene #12260) and pMD2.G (Addgene #12259) using Lipofectamine 3000 (Invitrogen) transfection reagent as per manufacturer instructions. After overnight incubation, medium was removed, and fresh medium added containing 5% serum and 1X Pen-strep. Cells were further incubated for 48h and 72h post-transfection and the virus containing supernatant collected and centrifuged for 10 mins at 500g to remove any cell debris. The supernatant was concentrated using Lentivirus Precipitation Solution (cat. #VC100; AlStem) according to the manufacturer's instructions and used to transduce SH-SY5Y cells. SH-SY5Y cell line was transduced with virus in complete medium (DMEM with 10% serum and 1X Pen-Strep) containing 8 μg/ml Polybrene (Sigma) with spin infection. Medium was changed 3-4 times to remove any virus residues and cells were used for further assays.

### qPCR analysis

Total RNA and DNA were extracted using AllPrep DNA/RNA Kits (Qiagen) according to the manufacture instruction. RNA and DNA concentrations and quality were verified by QIAxpert (Qiagen). Reverse transcription of RNA was performed using the manufacturer's instructions of iScript™ cDNA Synthesis Kit (Bio-Rad Laboratories, Inc., CA) with OligodT and Random Hexamer primers, generating cDNA for subsequent gene expression analysis. Real-time PCR was performed on a BioRad CFX384 Touch Real-Time PCR Detection System using iQ™ SYBR® Green Supermix (Bio-Rad Laboratories, Inc., CA) in a three step PCR: 95 °C:3 min, followed by 40 cycles of 95 °C:10 s, 60 °C:30 s and 72 °C:30 s. The relative expressions of the selected genes were normalized to the reference gene *GAPDH*. The specificity of real time PCR reaction was verified by the melt curve analysis. The expression level of selected genes were analyzed using ΔΔCT method (16) and compared via independent t-test. All statistical analyses were performed in GraphPad Prism 6 and 7 (GraphPad Software).

### Correlation network analysis in MS brain

*Raw data analysis.* The fastq files corresponding to bulk gene expression (RNA-sequencing) data from brain tissue samples of progressive MS patients (n = 12) and non-neurological controls (n = 10) (3, 4) were extracted from the RAW RNA sequence files and checked for quality control using multiqc software to make them ready for alignment (17). After trimming using the trimgalore program (18), fastqc files were aligned and annotated using STAR aligner and Stringtie software (19) by applying human hg38 refseq information from UCSC. The analysis was performed on the extracted count matrix using bash and Python.

*Network analysis.* In order to utilize a brain-specific network module, we applied a previously established bioinformatic pipeline utilizing co-expression network analysis (20), as briefly described below. The count matrix was loaded into R(3.6.1) environment and quantile normalized using the glimma package (21). Spearman correlation is applied on every gene pair and permuted for 10000 times to determine which interactions are significant (FDR < 0.05), which avoids biased filtering of the network based on correlation R value. The function also integrates hub connectivity significance for including only the interactions that have significant connectivity in the network. The produced network consisted of 5 million interactions among 27,059 genes, which limits the inherent resolution for defining modules which overlooks the multiscale organization of the network where compact clusters co-exist. In order to overcome this limitation, the correlation network was embedded on a spherical surface, thereby creating a planar maximally filtered network devoid of cross links. The final network consisted of 0.5 million interactions among 27,059 expressed genes from the RNAseq data. The planar maximally filtered graph is then clustered by implementing multiscale clustering algorithm (MCA) from the MEGENA package in R. MCA incorporates three distinct criteria to identify locally coherent clusters while maintaining a globally optimal partition. First, shortest path distances are utilized to optimize within-cluster compactness. Second, local path index is used to optimize local clustering structure. Third, overall modularity is employed to identify optimal partition. The final network clustered into 757 non-overlapping modules.

For the validation data, we applied the same pipeline as described above on several datasets (SI Appendix Table S11). In the $CUX2^+$ neuronal snRNA-seq count data (4), planar maximal filtration of the Spearmen correlated network of 10780 genes was multiscale clustered using the MEGENA package in R. This resulted in 91 modules out of which 1 module with *CHD1L* was significant and was further analyzed with Fisher enrichment test and pathway analysis using clusterProfiler.

*Cluster Trait association analysis.* Principal component analysis (PCA) is first performed for each cluster. Next, correlation between the first principal component and each trait was computed as cluster relevance to the trait. The 757 clusters identified from the correlation network were evaluated for the relevance to PPMS, SPMS and control phenotypes. Three clustered passed FDR P-value < 0.05.

### Figures S1 to S8

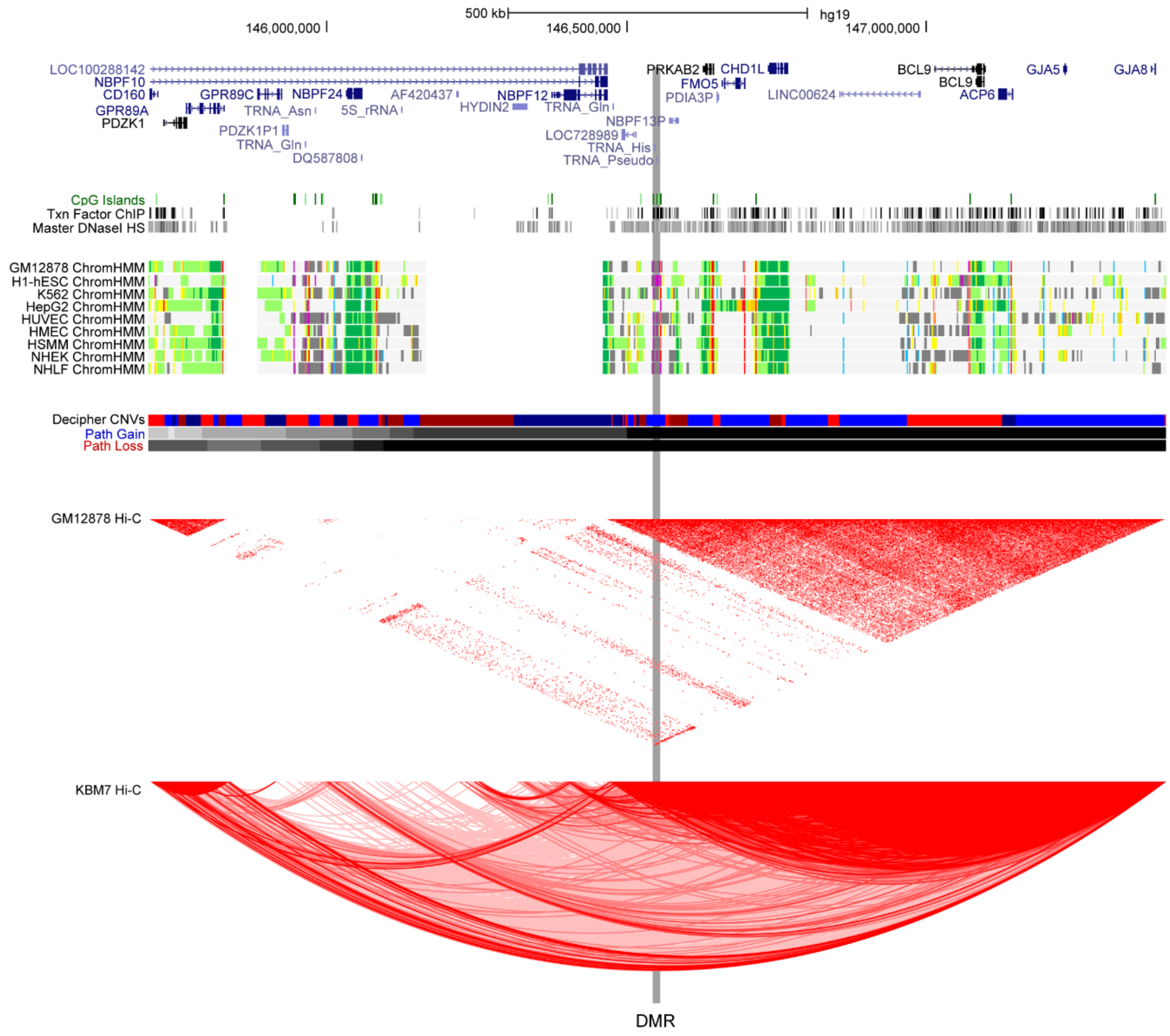


**Fig. S1.** Chromatin chromosome interaction (Hi-C) annotation of the locus from Roadmap. The Hi-C annotation showing chromatin folding data from in-situ experiments on several cell lines(22) displayed by UCSC genome browser. The data indicate how many interactions were detected between regions of the genome. Two cell lines were chosen as examples: the color shade in the triangle mode shows the proximity score for two genomic regions, intersection was further depicted using arcs drawn between the centers of interacting regions. A high score between two regions suggests that they are probably in close proximity in 3D space within the nucleus of a cell.

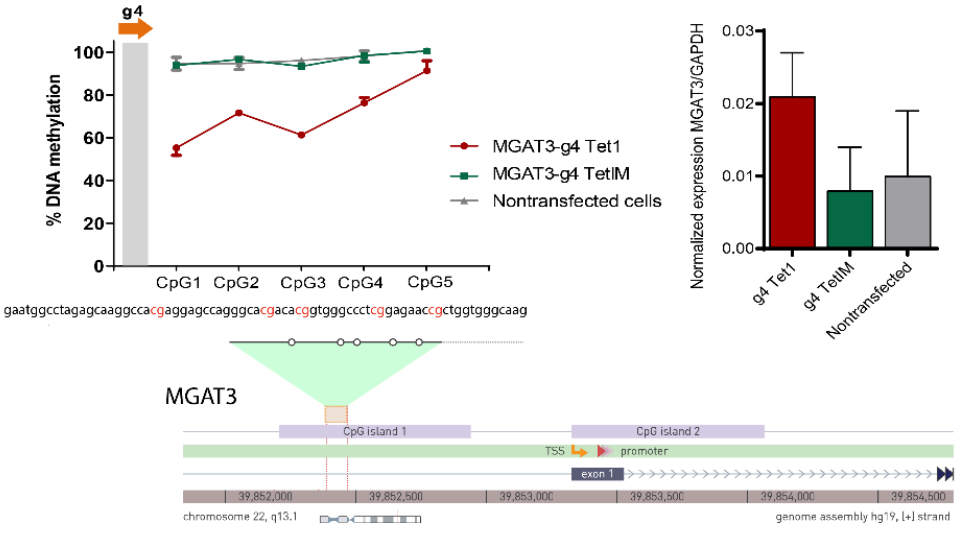


**Fig. S2.** Validation of the editing efficiency of dCas9-TET1 constructs by targeting 5 CpGs in the *MGAT3* gene promoter. The methylation decreased 10-50% in different CpGs, resulting in no significant changes in MGAT3 gene expression, as previously described(23).

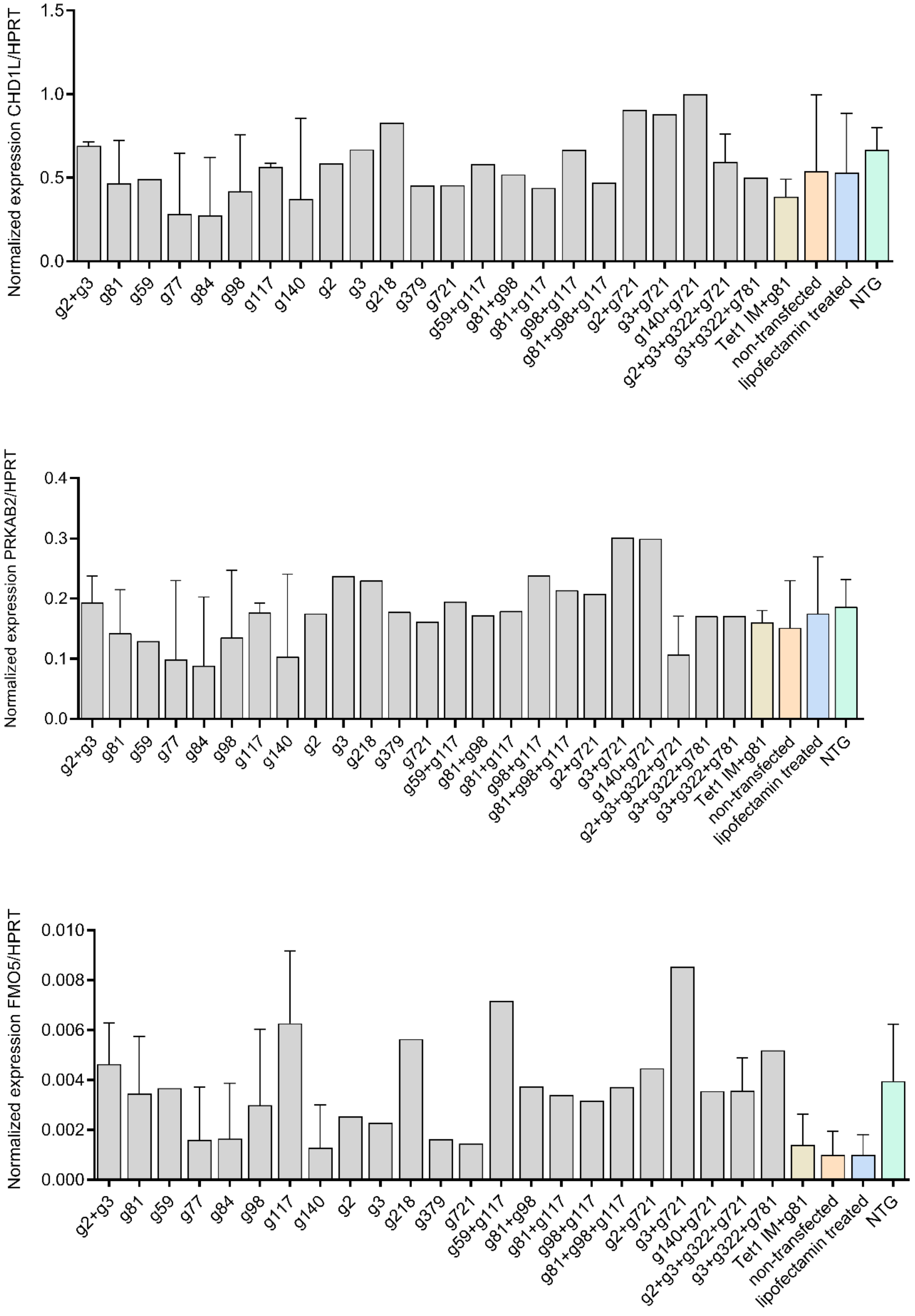


**Fig. S3.** Expression of the selected genes in the 1q21.1 locus in HEK293T cells. Results represent the relative expression normalized to HPRT (± SD).

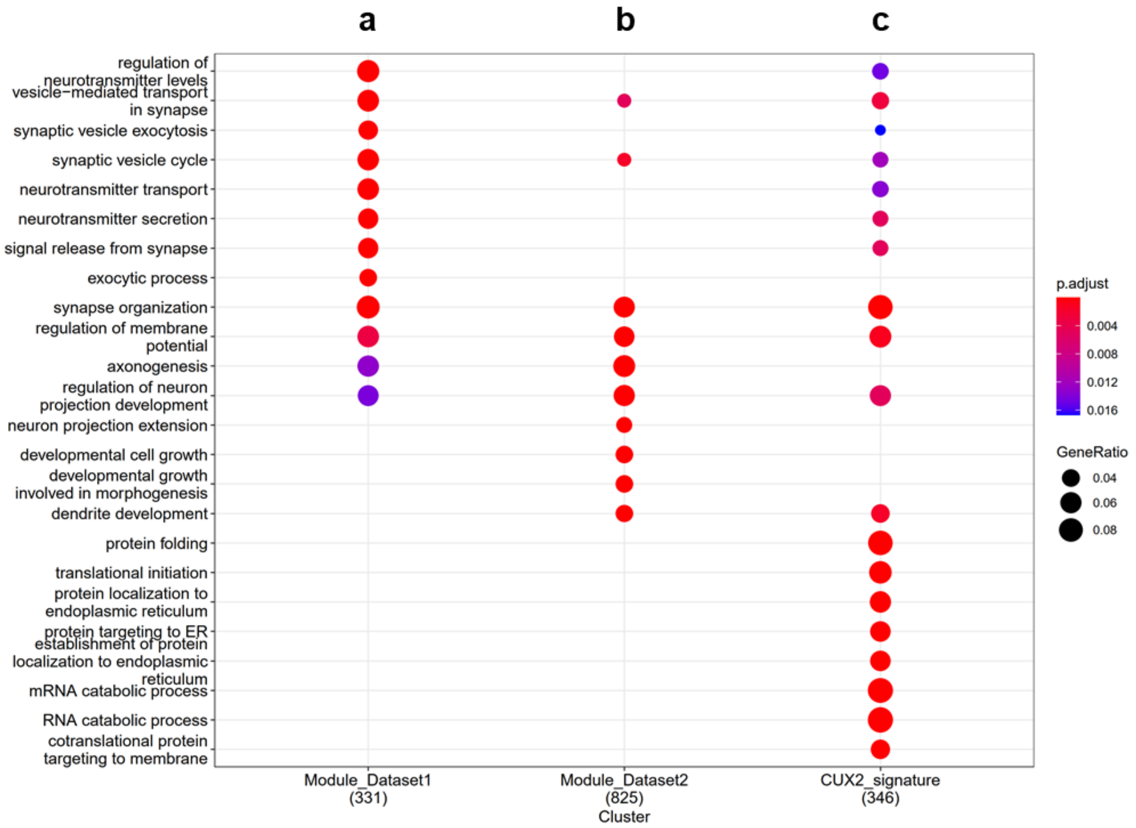


**Fig. S4.** Enrichment dot plot showing Gene Ontology (GO): Biological processes for **a.** *CHD1L* module from our study. **b.** Validated module from snRNA-seq data (PRJNA544731) **c.** gene set for CUX2+ neuronal gene signature. The size of the dot corresponds to the gene ratio overlapping with the pathway and the color of the dot represents significance of the FDR p-value of the enrichment.

**>1q21.1 DMR_1st (CpG 1&2 in 450K) [285 bps]**
GGGATGTTAAGGTTTGAGTTTTTTATGATATTCGGGGTGTTTCGATTTTATTTTGGGAGTTTTT
TCGTTTTTGTTTGTTTTTGGTTATTTAGTGCGCGGTTTTGGTTCGGTGATTCGTTATTGTTTTT
TTATGCGTTGTCGTTTTATAAAGGCGTATTTTTAGGTCGGTGGTTAGGTTTTGTTCGGACGTT
GTAATTGGTTTCGGGATTTGTGTTTTTGGTGAGTTTAGCGGAGTGTTGGGTGCGTCGTTTGC
GTGTTTTTTTTTTTGGAGAAAGGAGGGAGGGAA

**>1q21.1 DMR_2st (CpG 3 in 450K) [265 bps]**
GTTGTAATTGGTTTCGGGATTTGTGTTTTTGGTGAGTTTAGCGGAGTGTTGGGTGCGTCGTT
TGCGTGTTTTTTTTTTTGGAGAAAGGAGGGAGGGAACGGTTTTGTGAGACGATTTTAGGAGCG
ATTAGCGATTTTTATAAGTTTTAAGTTTTTTTAGCGTATAGGGAAATTGTTATTTATAGGGAAA
TTGTCGTTTTAGTGGTAAGAAGGTTGAAAAAGTTTTTTTTGTGTTTTTTTGGTGGTTTGGTGGT
TAGAATTTAGCGT

**>1q21.1 DMR_3st (CpG 4 in 450K) [131 bps]**
GGTAATTTTTGGTGGGTTATTTTTTATGATATAGCGTTTTTATTTATTTTCGTGTTTGTTATTCG
TATAAGCGGTTTTAGAGAGCGATTGAGCGTTTCGTTTAGGTGTATATCGTTGTGTAGAGATGT
TAG

**>MGAT3 [118 bps]**
GTTTTTGAGTTTTGAGAGGAATGGTTTAGAGTAAGGTTACGAGGAGTTAGGGTACGATACGG
TGGGTTTTCGGAGAATCGTTGGTGGGTAAGTGGTAGGAGAGTAGGTTTAAGAGGGT

**Fig. S5.** Bisulfite amplicons for the pyrosequencing assays. CpG sites are highlighted in yellow. Sequencing primers are underlined.

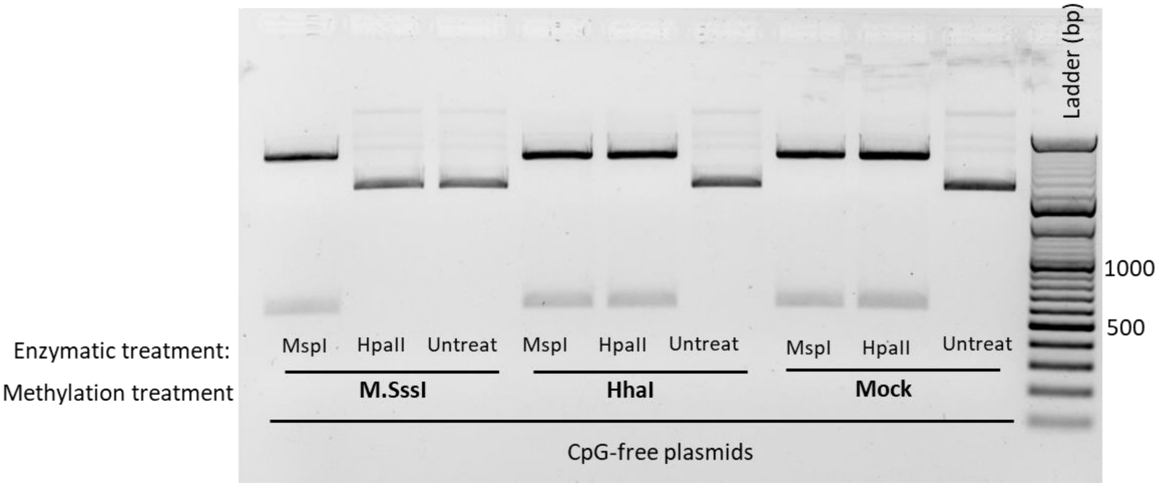


**Fig. S6.** Methylation of 1q21.1 DMR vector used for in-vitro DNA methylation assay. Representative gel of completely methylated (57 CpGs) by *M.SssI* or partially methylated (7 CpGs residing in the GCGC sequence) by *HhaI* methyltransferases 1q21.1 DMR containing vector and empty constructs, treated with the methyl-insensitive MspI or the methyl-sensitive HpaII. The mock control was treated equally but in absence of any methyltransferases and correspond to unmethylated inserts.

**P3-dCas9-Tet1-GFP-Puro (Addgene #190728)**

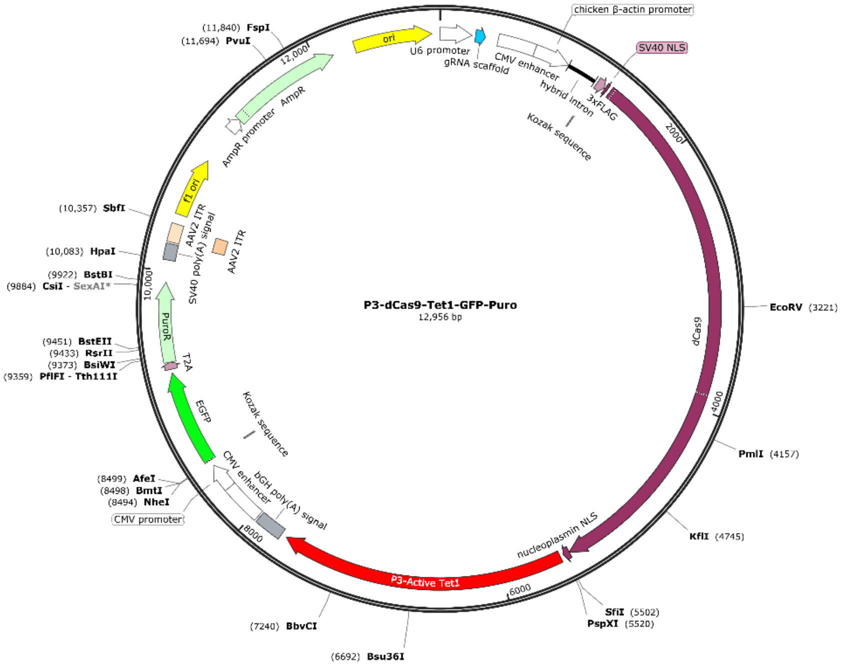


GGCCgcctatttcccatgattccttcatatttgcatatacgatacaaggctgttagagagataattggaattaatttgactgtaaacacaaa
gatattagtacaaaatacgtgacgtagaaagtaataatttcttgggtagtttgcagttttaaaattatgttttaaaatggactatcatatgcttac
cgtaacttgaaagtatttcgatttcttggctttatatatcttgtggaaaggacgaaacaccgggtcttcgagaagacctgttttagagctagaa
atagcaagttaaaataaggctagtccgttatcaacttgaaaaagtggcaccgagtcggtgctttttttgttttagagctagaaatagcaagtta
aaataaggctagtccgtttttagcgcgtgcgccaattctgcagacaaatggctctagaggtacccgttacataacttacggtaaatggccc
gcctggctgaccgcccaacgacccccgcccattgacgtcaatagtaacgccaatagggactttccattgacgtcaatgggtggagtattt
acggtaaactgcccacttggcagtacatcaagtgtatcatatgccaagtacgccccctattgacgtcaatgacggtaaatggcccgcctg
gcattgtgcccagtacatgaccttatgggactttcctacttggcagtacatctacgtattagtcatcgctattaccatggtcgaggtgagcccc
acgttctgcttcactctccccatctcccccccctccccacccccaattttgtatttatttattttttaattattttgtgcagcgatgggggcgggggg
gggggggggcgcgcgccaggcggggcggggcggggcgaggggcggggcggggcgaggcggagaggtgcggcggcagcc
aatcagagcggcgcgctccgaaagtttccttttatggcgaggcggcggcggcggcggccctataaaaagcgaagcgcgcggcgggc
gggagtcgctgcgacgctgccttcgccccgtgccccgctccgccgccgcctcgcgccgcccgccccggctctgactgaccgcgttactc
ccacaggtgagcgggcgggacggcccttctcctccgggctgtaattagctgagcaagaggtaagggtttaagggatggttggttggtgg
ggtattaatgtttaattacctggagcacctgcctgaaatcactttttttcaggttggaccggtgccaccatggactataaggaccacgacgg
agactacaaggatcatgatattgattacaaagacgatgacgataagatggccccaaagaagaagcggaaggtcggtatccacggag
tcccagcagccgacaagaagtacagcatcggcctggccatcggcaccaactctgtgggctgggccgtgatcaccgacgagtacaag
gtgcccagcaagaaattcaaggtgctgggcaacaccgaccggcacagcatcaagaagaacctgatcggagccctgctgttcgacag
cggcgaaacagccgaggccacccggctgaagagaaccgccagaagaagatacaccagacggaagaaccggatctgctatctgc
aagagatcttcagcaacgagatggccaaggtggacgacagcttcttccacagactggaagagtccttcctggtggaagaggataaga
agcacgagcggcaccccatcttcggcaacatcgtggacgaggtggcctaccacgagaagtaccccaccatctaccacctgagaaag
aaactggtggacagcaccgacaaggccgacctgcggctgatctatctggccctggcccacatgatcaagttccggggccacttcctgat
cgagggcgacctgaaccccgacaacagcgacgtggacaagctgttcatccagctggtgcagacctacaaccagctgttcgaggaaa
accccatcaacgccagcggcgtggacgccaaggccatcctgtctgccagactgagcaagagcagacggctggaaaatctgatcgcc
cagctgcccggcgagaagaagaatggcctgttcggcaacctgattgccctgagcctgggcctgacccccaacttcaagagcaacttcg
acctggccgaggatgccaaactgcagctgagcaaggacacctacgacgacgacctggacaacctgctggcccagatcggcgacca
gtacgccgacctgtttctggccgccaagaacctgtccgacgccatcctgctgagcgacatcctgagagtgaacaccgagatcaccaag
gccccccctgagcgcctctatgatcaagagatacgacgagcaccaccaggacctgaccctgctgaaagctctcgtgcggcagcagctg
cctgagaagtacaaagagattttcttcgaccagagcaagaacggctacgccggctacattgacggcggagccagccaggaagagtt
ctacaagttcatcaagcccatcctggaaaagatggacggcaccgaggaactgctcgtgaagctgaacagagaggacctgctgcgga
agcagcggaccttcgacaacggcagcatcccccaccagatccacctgggagagctgcacgccattctgcggcggcaggaagattttt
acccattcctgaaggacaaccgggaaaagatcgagaagatcctgaccttccgcatcccctactacgtgggccctctggccaggggaa

acagcagattcgcctggatgaccagaaagagcgaggaaaccatcacccccctggaacttcgaggaagtggtggacaagggcgcttcc
gcccagagcttcatcgagcggatgaccaacttcgataagaacctgcccaacgagaaggtgctgcccaagcacagcctgctgtacga
gtacttcaccgtgtataacgagctgaccaaagtgaaatacgtgaccgagggaatgagaaagcccgccttcctgagcggcgagcaga
aaaaggccatcgtggacctgctgttcaagaccaaccggaaagtgaccgtgaagcagctgaaagaggactacttcaagaaaatcgag
tgcttcgactccgtggaaatctccggcgtggaagatcggttcaacgcctccctgggcacataccacgatctgctgaaaattatcaaggac
aaggacttcctggacaatgaggaaaacgaggacattctggaagatatcgtgctgaccctgacactgtttgaggacagagagatgatcg
aggaacggctgaaaacctatgcccacctgttcgacgacaaagtgatgaagcagctgaagcggcggagatacaccggctggggcag
gctgagccggaagctgatcaacggcatccgggacaagcagtccggcaagacaatcctggatttcctgaagtccgacggcttcgccaa
cagaaacttcatgcagctgatccacgacgacagcctgacctttaaagaggacatccagaaagcccaggtgtccggccagggcgata
gcctgcacgagcacattgccaatctggccggcagccccgccattaagaagggcatcctgcagacagtgaaggtggtggacgagctc
gtgaaagtgatgggccggcacaagcccgagaacatcgtgatcgaaatggccagagagaaccagaccacccagaagggacagaa
gaacagccgcgagagaatgaagcggatcgaagagggcatcaaagagctgggcagccagatcctgaaagaacaccccgtggaaa
acacccagctgcagaacgagaagctgtacctgtactacctgcagaatgggcgggatatgtacgtggaccaggaactggacatcaacc
ggctgtccgactacgatgtggacgctatcgtgcctcagagctttctgaaggacgactccatcgacaacaaggtgctgaccagaagcga
caagaaccggggcaagagcgacaacgtgccctccgaagaggtcgtgaagaagatgaagaactactggcggcagctgctgaacgc
caagctgattacccagagaaagttcgacaatctgaccaaggccgagagaggcggcctgagcgaactggataaggccggcttcatca
agagacagctggtggaaacccggcagatcacaaagcacgtggcacagatcctggactcccggatgaacactaagtacgacgaga
atgacaagctgatccgggaagtgaaagtgatcaccctgaagtccaagctggtgtccgatttccggaaggatttccagttttacaaagtgc
gcgagatcaacaactaccaccacgcccacgacgcctacctgaacgccgtcgtgggaaccgccctgatcaaaaagtaccctaagctg
gaaagcgagttcgtgtacggcgactacaaggtgtacgacgtgcggaagatgatcgccaagagcgagcaggaaatcggcaaggcta
ccgccaagtacttcttctacagcaacatcatgaactttttcaagaccgagattaccctggccaacggcgagatccggaagcggcctctga
tcgagacaaacggcgaaaccggggagatcgtgtgggataagggccgggattttgccaccgtgcggaaagtgctgagcatgccccaa
gtgaatatcgtgaaaaagaccgaggtgcagacaggcggcttcagcaaagagtctatcctgcccaagaggaacagcgataagctgat
cgccagaaagaaggactgggaccctaagaagtacggcggcttcgacagccccaccgtggcctattctgtgctggtggtggccaaagt
ggaaaagggcaagtccaagaaactgaagagtgtgaaagagctgctggggatcaccatcatggaaagaagcagcttcgagaagaa
tcccatcgactttctggaagccaagggctacaaagaagtgaaaaaggacctgatcatcaagctgcctaagtactccctgttcgagctgg
aaaacggccggaagagaatgctggcctctgccggcgaactgcagaagggaaacgaactggccctgccctccaaatatgtgaacttc
ctgtacctggccagccactatgagaagctgaagggctcccccgaggataatgagcagaaacagctgtttgtggaacagcacaagcac
tacctggacgagatcatcgagcagatcagcgagttctccaagagagtgatcctggccgacgctaatctggacaaagtgctgtccgccta
caacaagcaccgggataagcccatcagagagcaggccgagaatatcatccacctgtttaccctgaccaatctgggagcccctgccgc
cttcaagtactttgacaccaccatcgaccggaagaggtacaccagcaccaaagaggtgctggacgccaccctgatccaccagagcat
caccggcctgtacgagacacggatcgacctgtctcagctgggaggcgacaaaaggccggcggccacgaaaaaggccggacagg
ccaaaaagaaaaagctcgagggcggaggcgggagcggatccctgcccacctgcagctgtcttgatcgagttatacaaaaagacaa
aggcccatattatacacaccttggggcaggaccaagtgttgctgctgtcagggaaatcatggagaataggtatggtcaaaaaggaaac
gcaataaggatagaaatagtagtgtacaccggtaaagaagggaaaagctctcatgggtgtccaattgctaagtgggttttaagaagaa
gcagtgatgaagaaaaagttctttgtttggtccggcagcgtacaggccaccactgtccaactgctgtgatggtggtgctcatcatggtgtgg
gatggcatccctcttccaatggccgaccggctatacacagagctcacagagaatctaaagtcatacaatgggcaccctaccgacaga
agatgcaccctcaatgaaaatcgtacctgtacatgtcaaggaattgatccagagacttgtggagcttcattctcttttggctgttcatggagta
tgtactttaatggctgtaagtttggtagaagcccaagccccagaagatttagaattgatccaagctctcccttacatgaaaaaaaccttgaa
gataacttacagagtttggctacacgattagctccaatttataagcagtatgctccagtagcttaccaaaatcaggtggaatatgaaaatgt
tgcccgagaatgtcggcttggcagcaaggaaggtcgacccttctctggggtcactgcttgcctggacttctgtgctcatccccacagggac
attcacaacatgaataatggaagcactgtggtttgtaccttaactcgagaagataaccgctctttgggtgttattcctcaagatgagcagctc
catgtgctacctctttataagctttcagacacagatgagtttggctccaaggaaggaatggaagccaagatcaaatctggggccatcgag
gtcctggcaccccgccgcaaaaaaagaacgtgtttcactcagcctgttccccgttctggaaagaagagggctgcgatgatgacagagg
ttcttgcacataagataagggcagtggaaaagaaacctattccccgaatcaagcggaagaataactcaacaacaacaaacaacagt
aagccttcgtcactgccaaccttagggagtaacactgagaccgtgcaacctgaagtaaaaagtgaaaccgaaccccattttatcttaaa
aagttcagacaacactaaaacttattcgctgatgccatccgctcctcacccagtgaaagaggcatctccaggcttctcctggtcccccaa
gactgcttcagccacaccagctccactgaagaatgacgcaacagcctcatgcgggttttcagaaagaagcagcactccccactgtacg
atgccttcgggtagactcagtggtgccaatgctgcagctgctgatggccctggcatttcacagcttggcgaagtggctcctctccccaccct
gtctgctcctgtgatggagcccctcattaattctgagccttccactggtgtgactgagccgctaacgcctcatcagccaaaccaccagccct
ccttcctcacctctcctcaagaccttgcctcttctccaatggaagaagatgagcagcattctgaagcagatgagcctccatcagacgaac
ccctatctgatgacccccctgtcacctgctgaggagaaattgccccacattgatgagtattggtcagacagtgagcacatctttttggatgca
aatattggtggggtggccatcgcacctgctcacggctcggttttgattgagtgtgcccggcgagagctgcacgctaccactcctgttgagc
accccaaccgtaatcatccaacccgcctctcccttgtcttttaccagcacaaaaacctaaataagccccaacatggttttgaactaaaca
agattaagtttgaggctaaagaagctaagaataagaaaatgaaggcctcagagcaaaaagaccaggcagctaatgaaggtccaga
acagtcctctgaagtaaatgaattgaaccaaattccttctcataaagcattaacattaacccatgacaatgttgtcaccgtgtccccttatgct

ctcacacacgttgcggggccctataaccattgggtctgagaattcaattctaactagagctcgctgatcagcctcgactgtgccttctagttg
ccagccatctgttgtttgcccctcccccgtgccttccttgaccctggaaggtgccactcccactgtcctttcctaataaaatgaggaaattgca
tcgcattgtctgagtaggtgtcattctattctggggggtggggtggggcaggacagcaagggggaggattgggaagagaatagcaggc
atgctggggagcggccgcgttccgcgttacataacttacggtaaatggcccgcctggctgaccgcccaacgacccccgcccattgacg
tcaataatgacgtatgttcccatagtaacgccaatagggactttccattgacgtcaatgggtggagtatttacggtaaactgcccacttggc
agtacatcaagtgtatcatatgccaagtacgccccctattgacgtcaatgacggtaaatggcccgcctggcattatgcccagtacatgac
cttatgggactttcctacttggcagtacatctacgtattagtcatcgctattaccatggtgatgcggttttggcagtacatcaatgggcgtggat
agcggtttgactcacggggatttccaagtctccaccccattgacgtcaatgggagtttgttttggcaccaaaatcaacgggactttccaaaa
tgtcgtaacaactccgccccattgacgcaaatgggcggtaggcgtgtacggtgggaggtctatataagcagagctggtttagtgaaccgt
cagatccgctagcgctaccggtcgccaccatggtgagcaagggcgaggagctgttcaccggggtggtgcccatcctggtcgagctgg
acggcgacgtaaacggccacaagttcagcgtgtccggcgagggcgagggcgatgccacctacggcaagctgaccctgaagttcatc
tgcaccaccggcaagctgcccgtgccctggcccaccctcgtgaccaccctgacctacggcgtgcagtgcttcagccgctaccccgacc
acatgaagcagcacgacttcttcaagtccgccatgcccgaaggctacgtccaggagcgcaccatcttcttcaaggacgacggcaacta
caagacccgcgccgaggtgaagttcgagggcgacaccctggtgaaccgcatcgagctgaagggcatcgacttcaaggaggacggc
aacatcctggggcacaagctggagtacaactacaacagccacaacgtctatatcatggccgacaagcagaagaacggcatcaaggt
gaacttcaagatccgccacaacatcgaggacggcagcgtgcagctcgccgaccactaccagcagaacacccccatcggcgacggc
cccgtgctgctgcccgacaaccactacctgagcacccagtccgccctgagcaaagaccccaacgagaagcgcgatcacatggtcct
gctggagttcgtgaccgccgccgggatcactctcggcatggacgagctgtacaagtccggactcagatctcgagctcaagcttctgagg
gcagaggaagtctgctaacatgcggtgacgtcgaggagaatcctggcccaatgaccgagtacaagcccacggtgcgcctcgccacc
cgcgacgacgtccccagggccgtacgcaccctcgccgccgcgttcgccgactaccccgccacgcgccacaccgtcgatccggaccg
ccacatcgagcgggtcaccgagctgcaagaactcttcctcacgcgcgtcgggctcgacatcggcaaggtgtgggtcgcggacgacgg
cgccgcggtggcggtctggaccacgccggagagcgtcgaagcgggggcggtgttcgccgagatcggcccgcgcatggccgagttg
agcggttcccggctggccgcgcagcaacagatggaaggcctcctggcgccgcaccggcccaaggagcccgcgtggttcctggccac
cgtcggcgtctcgcccgaccaccagggcaagggtctgggcagcgccgtcgtgctccccggagtggaggcggccgagcgcgccggg
gtgcccgccttcctggagacctccgcgccccacaacctccccttctacgagcggctcggcttcaccgtcaccgccgacgtcgaggtgcc
cgaaggaccgcgcacctggtgcatgacccgcaagcccggtgcctgaaagcttcgaattctgcagtcgacggtaccgcgggcccggg
atccaccggatctagataactgatcataatcagccataccacatttgtagaggttttacttgctttaaaaaacctcccacacctccccctgaa
cctgaaacataaaatgaatgcaattgttgttgttaacttgtttattgcagcttataatggttacaaataaagcaatagcatcacaaatttcaca
aataaagcatttttttcactgcattctagttgtggtttgtccaaactcatcaatgtatcttaacgcggcggccgcaggaacccctagtgatgga
gttggccactccctctctgcgcgctcgctcgctcactgaggccgggcgaccaaaggtcgcccgacgcccgggctttgcccgggcggcc
tcagtgagcgagcgagcgcgcagctgcctgcaggggcgcctgatgcggtattttctccttacgcatctgtgcggtatttcacaccgcatac
gtcaaagcaaccatagtacgcgccctgtagcggcgcattaagcgcggcgggtgtggtggttacgcgcagcgtgaccgctacacttgcc
agcgccctagcgcccgctcctttcgctttcttcccttcctttctcgccacgttcgccggctttccccgtcaagctctaaatcgggggctcccttta
gggttccgatttagtgctttacggcacctcgaccccaaaaaacttgatttgggtgatggttcacgtagtgggccatcgccctgatagacggt
ttttcgccctttgacgttggagtccacgttctttaatagtggactcttgttccaaactggaacaacactcaaccctatctcgggctattcttttgatt
tataagggattttgccgatttcggcctattggttaaaaaatgagctgatttaacaaaaatttaacgcgaattttaacaaaatattaacgtttac
aattttatggtgcactctcagtacaatctgctctgatgccgcatagttaagccagccccgacacccgccaacacccgctgacgcgccctg
acgggcttgtctgctcccggcatccgcttacagacaagctgtgaccgtctccgggagctgcatgtgtcagaggttttcaccgtcatcaccg
aaacgcgcgagacgaaagggcctcgtgatacgcctatttttataggttaatgtcatgataataatggtttcttagacgtcaggtggcactttt
cggggaaatgtgcgcggaacccctatttgtttatttttctaaatacattcaaatatgtatccgctcatgagacaataaccctgataaatgcttc
aataatattgaaaaaggaagagtatgagtattcaacatttccgtgtcgcccttattcccttttttgcggcattttgccttcctgtttttgctcaccca
gaaacgctggtgaaagtaaaagatgctgaagatcagttgggtgcacgagtgggttacatcgaactggatctcaacagcggtaagatcc
ttgagagttttcgccccgaagaacgttttccaatgatgagcacttttaaagttctgctatgtggcgcggtattatcccgtattgacgccgggca
agagcaactcggtcgccgcatacactattctcagaatgacttggttgagtactcaccagtcacagaaaagcatcttacggatggcatgac
agtaagagaattatgcagtgctgccataaccatgagtgataacactgcggccaacttacttctgacaacgatcggaggaccgaaggag
ctaaccgcttttttgcacaacatgggggatcatgtaactcgccttgatcgttgggaaccggagctgaatgaagccataccaaacgacga
gcgtgacaccacgatgcctgtagcaatggcaacaacgttgcgcaaactattaactggcgaactacttactctagcttcccggcaacaatt
aatagactggatggaggcggataaagttgcaggaccacttctgcgctcggcccttccggctggctggtttattgctgataaatctggagcc
ggtgagcgtggaagccgcggtatcattgcagcactggggccagatggtaagccctcccgtatcgtagttatctacacgacggggagtc
aggcaactatggatgaacgaaatagacagatcgctgagataggtgcctcactgattaagcattggtaactgtcagaccaagtttactcat
atatactttagattgatttaaaacttcatttttaatttaaaaggatctaggtgaagatcctttttgataatctcatgaccaaaatcccttaacgtga
gttttcgttccactgagcgtcagaccccgtagaaaagatcaaaggatcttcttgagatcctttttttctgcgcgtaatctgctgcttgcaaacaa
aaaaaccaccgctaccagcggtggtttgtttgccggatcaagagctaccaactctttttccgaaggtaactggcttcagcagagcgcaga
taccaaatactgtccttctagtgtagccgtagttaggccaccacttcaagaactctgtagcaccgcctacatacctcgctctgctaatcctgtt
accagtggctgctgccagtggcgataagtcgtgtcttaccgggttggactcaagacgatagttaccggataaggcgcagcggtcgggct
gaacggggggttcgtgcacacagcccagcttggagcgaacgacctacaccgaactgagatacctacagcgtgagctatgagaaagc

gccacgcttcccgaagggagaaaggcggacaggtatccggtaagcggcagggtcggaacaggagagcgcacgagggagcttcc
agggggaaacgcctggtatctttatagtcctgtcgggtttcgccacctctgacttgagcgtcgatttttgtgatgctcgtcaggggggcggag
cctatggaaaaacgccagcaacgcggcctttttacggttcctggccttttgctggccttttgctcacatgt

**P3-Lenti-dCas9-Tet1-GFP (Addgene #190729)**

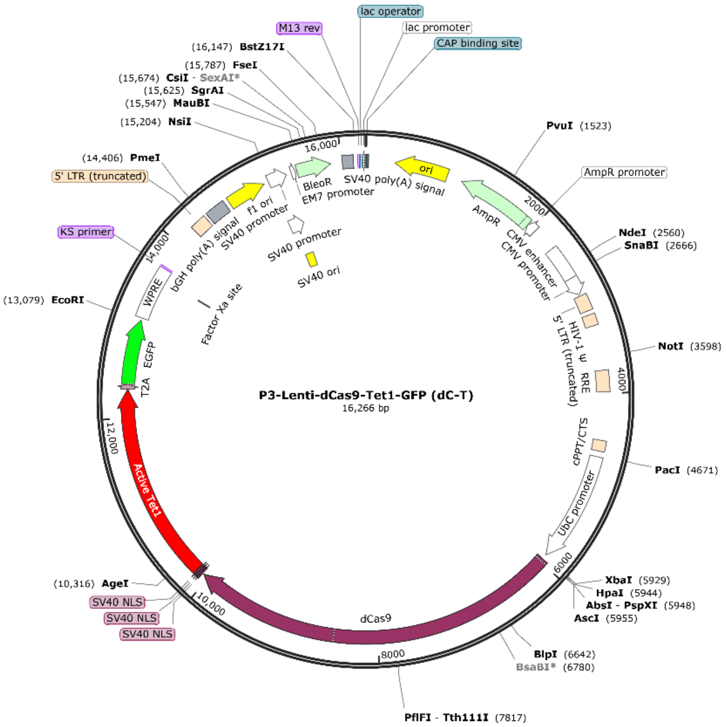


tggggtgcctaatgagtgagctaactcacattaattgcgttgcgctcactgcccgctttccagtcgggaaacctgtcgtgccagctgcatta
atgaatcggccaacgcgcggggagaggcggtttgcgtattgggcgctcttccgcttcctcgctcactgactcgctgcgctcggtcgttcgg
ctgcggcgagcggtatcagctcactcaaaggcggtaatacggttatccacagaatcaggggataacgcaggaaagaacatgtgagc
aaaaggccagcaaaaggccaggaaccgtaaaaaggccgcgttgctggcgtttttccataggctccgcccccctgacgagcatcaca
aaaatcgacgctcaagtcagaggtggcgaaacccgacaggactataaagataccaggcgtttccccctggaagctccctcgtgcgct
ctcctgttccgaccctgccgcttaccggatacctgtccgcctttctcccttcgggaagcgtggcgctttctcatagctcacgctgtaggtatctc
agttcggtgtaggtcgttcgctccaagctgggctgtgtgcacgaaccccccgttcagcccgaccgctgcgccttatccggtaactatcgtct
tgagtccaacccggtaagacacgacttatcgccactggcagcagccactggtaacaggattagcagagcgaggtatgtaggcggtgc
tacagagttcttgaagtggtggcctaactacggctacactagaagaacagtatttggtatctgcgctctgctgaagccagttaccttcggaa
aaagagttggtagctcttgatccggcaaacaaaccaccgctggtagcggtggtttttttgtttgcaagcagcagattacgcgcagaaaaa
aaggatctcaagaagatcctttgatcttttctacggggtctgacgctcagtggaacgaaaactcacgttaagggattttggtcatgagattat
caaaaaggatcttcacctagatccttttaaattaaaaatgaagttttaaatcaatctaaagtatatatgagtaaacttggtctgacagttacca
atgcttaatcagtgaggcacctatctcagcgatctgtctatttcgttcatccatagttgcctgactccccgtcgtgtagataactacgatacgg
gagggcttaccatctggccccagtgctgcaatgataccgcgagacccacgctcaccggctccagatttatcagcaataaaccagccag
ccggaagggccgagcgcagaagtggtcctgcaactttatccgcctccatccagtctattaattgttgccgggaagctagagtaagtagttc
gccagttaatagtttgcgcaacgttgttgccattgctacaggcatcgtggtgtcacgctcgtcgtttggtatggcttcattcagctccggttccc
aacgatcaaggcgagttacatgatcccccatgttgtgcaaaaaagcggttagctccttcggtcctccgatcgttgtcagaagtaagttggc
cgcagtgttatcactcatggttatggcagcactgcataattctcttactgtcatgccatccgtaagatgcttttctgtgactggtgagtactcaac
caagtcattctgagaatagtgtatgcggcgaccgagttgctcttgcccggcgtcaatacgggataataccgcgccacatagcagaacttt
aaaagtgctcatcattggaaaacgttcttcggggcgaaaactctcaaggatcttaccgctgttgagatccagttcgatgtaacccactcgt
gcacccaactgatcttcagcatcttttactttcaccagcgtttctgggtgagcaaaaacaggaaggcaaaatgccgcaaaaaagggaat
aagggcgacacggaaatgttgaatactcatactcttcctttttcaatattattgaagcatttatcagggttattgtctcatgagcggatacatatt
tgaatgtatttagaaaaataaacaaataggggttccgcgcacatttccccgaaaagtgccacctgacgtcgacggatcgggagatctcc
cgatcccctatggtgcactctcagtacaatctgctctgatgccgcatagttaagccagtatctgctccctgcttgtgtgttggaggtcgctgag
tagtgcgcgagcaaaatttaagctacaacaaggcaaggcttgaccgacaattgcatgaagaatctgcttagggttaggcgttttgcgctg
cttcgcgatgtacgggccagatatacgcgttgacattgattattgactagttattaatagtaatcaattacggggtcattagttcatagcccat
atatggagttccgcgttacataacttacggtaaatggcccgcctggctgaccgcccaacgacccccgcccattgacgtcaataatgacgt
atgttcccatagtaacgccaatagggactttccattgacgtcaatgggtggagtatttacggtaaactgcccacttggcagtacatcaagtg
tatcatatgccaagtacgccccctattgacgtcaatgacggtaaatggcccgcctggcattatgcccagtacatgaccttatgggactttcc
tacttggcagtacatctacgtattagtcatcgctattaccatggtgatgcggttttggcagtacatcaatgggcgtggatagcggtttgactca

cggggatttccaagtctccaccccattgacgtcaatgggagtttgttttggcaccaaaatcaacgggactttccaaaatgtcgtaacaactc
cgccccattgacgcaaatgggcggtaggcgtgtacggtgggaggtctatataagcagcgcgttttgcctgtactgggtctctctggttaga
ccagatctgagcctgggagctctctggctaactagggaacccactgcttaagcctcaataaagcttgccttgagtgcttcaagtagtgtgtg
cccgtctgttgtgtgactctggtaactagagatccctcagacccttttagtcagtgtggaaaatctctagcagtggcgcccgaacagggact
tgaaagcgaaagggaaaccagaggagctctctcgacgcaggactcggcttgctgaagcgcgcacggcaagaggcgaggggcgg
cgactggtgagtacgccaaaaattttgactagcggaggctagaaggagagagatgggtgcgagagcgtcagtattaagcgggggag
aattagatcgcgatgggaaaaaattcggttaaggccagggggaaagaaaaaatataaattaaaacatatagtatgggcaagcaggg
agctagaacgattcgcagttaatcctggcctgttagaaacatcagaaggctgtagacaaatactgggacagctacaaccatcccttcag
acaggatcagaagaacttagatcattatataatacagtagcaaccctctattgtgtgcatcaaaggatagagataaaagacaccaagg
aagctttagacaagatagaggaagagcaaaacaaaagtaagaccaccgcacagcaagcggccgctgatcttcagacctggagga
ggagatatgagggacaattggagaagtgaattatataaatataaagtagtaaaaattgaaccattaggagtagcacccaccaaggca
aagagaagagtggtgcagagagaaaaaagagcagtgggaataggagctttgttccttgggttcttgggagcagcaggaagcactatg
ggcgcagcgtcaatgacgctgacggtacaggccagacaattattgtctggtatagtgcagcagcagaacaatttgctgagggctattga
ggcgcaacagcatctgttgcaactcacagtctggggcatcaagcagctccaggcaagaatcctggctgtggaaagatacctaaagga
tcaacagctcctggggatttggggttgctctggaaaactcatttgcaccactgctgtgccttggaatgctagttggagtaataaatctctgga
acagatttggaatcacacgacctggatggagtgggacagagaaattaacaattacacaagcttaatacactccttaattgaagaatcgc
aaaaccagcaagaaaagaatgaacaagaattattggaattagataaatgggcaagtttgtggaattggtttaacataacaaattggctg
tggtatataaaattattcataatgatagtaggaggcttggtaggtttaagaatagtttttgctgtactttctatagtgaatagagttaggcaggg
atattcaccattatcgtttcagacccacctcccaaccccgaggggacccgacaggcccgaaggaatagaagaagaaggtggagaga
gagacagagacagatccattcgattagtgaacggatcggcactgcgtgcgccaattctgcagacaaatggcagtattcatccacaatttt
aaaagaaaaggggggattggggggtacagtgcaggggaaagaatagtagacataatagcaacagacatacaaactaaagaatta
caaaaacaaattacaaaaattcaaaattttcgggtttattacagggacagcagagatccagtttggttaattaacccgtgtcggctccaga
tctggcctccgcgccgggttttggcgcctcccgcgggcgcccccctcctcacggcgagcgctgccacgtcagacgaagggcgcagcg
agcgtcctgatccttccgcccggacgctcaggacagcggcccgctgctcataagactcggccttagaaccccagtatcagcagaagg
acattttaggacgggacttgggtgactctagggcactggttttctttccagagagcggaacaggcgaggaaaagtagtcccttctcggcg
attctgcggagggatctccgtggggcggtgaacgccgatgattatataaggacgcgccgggtgtggcacagctagttccgtcgcagcc
gggatttgggtcgcggttcttgtttgtggatcgctgtgatcgtcacttggtgagtagcgggctgctgggctggccggggctttcgtggccgcc
gggccgctcggtgggacggaagcgtgtggagagaccgccaagggctgtagtctgggtccgcgagcaaggttgccctgaactggggg
ttgggggggagcgcagcaaaatggcggctgttcccgagtcttgaatggaagacgcttgtgaggcgggctgtgaggtcgttgaaacaagg
tgggggggcatggtgggcggcaagaacccaaggtcttgaggccttcgctaatgcgggaaagctcttattcgggtgagatgggctggggc
accatctggggaccctgacgtgaagtttgtcactgactggagaactcggtttgtcgtctgttgcgggggcggcagttatggcggtgccgttg
ggcagtgcacccgtacctttgggagcgcgcgccctcgtcgtgtcgtgacgtcacccgttctgttggcttataatgcagggtggggccacct
gccggtaggtgtgcggtaggcttttctccgtcgcaggacgcagggttcgggcctagggtaggctctcctgaatcgacaggcgccggacc
tctggtgaggggagggataagtgaggcgtcagtttctttggtcggttttatgtacctatcttcttaagtagctgaagctccggttttgaactatgc
gctcggggttggcgagtgtgttttgtgaagttttttaggcaccttttgaaatgtaatcatttgggtcaatatgtaattttcagtgttagactagtaaa
ttgtccgctaaattctggccgtttttggcttttttgttagacgaagcttgggctgcaggtcgactctagaggatccagttaacctcgagggcgc
gcccatggacaagaagtattctatcggactggccatcgggactaatagcgtcgggtgggccgtgatcactgacgagtacaaggtgccc
tctaagaagttcaaggtgctcgggaacaccgaccggcattccatcaagaaaaatctgatcggagctctcctctttgattcaggggagacc
gctgaagcaacccgcctcaagcggactgctagacggcggtacaccaggaggaagaaccggatttgttaccttcaagagatattctcca
acgaaatggcaaaggtcgacgacagcttcttccataggctggaagaatcattcctcgtggaagaggataagaagcatgaacggcatc
ccatcttcggtaatatcgtcgacgaggtggcctatcacgagaaatacccaaccatctaccatcttcgcaaaaagctggtggactcaacc
gacaaggcagacctccggcttatctacctggccctggcccacatgatcaagttcagaggccacttcctgatcgagggcgacctcaatcc
tgacaatagcgatgtggataaactgttcatccagctggtgcagacttacaaccagctctttgaagagaaccccatcaatgcaagcggag
tcgatgccaaggccattctgtcagcccggctgtcaaagagccgcagacttgagaatcttatcgctcagctgccgggtgaaaagaaaaat
ggactgttcgggaacctgattgctctttcacttgggctgactcccaatttcaagtctaatttcgacctggcagaggatgccaagctgcaactg
tccaaggacacctatgatgacgatctcgacaacctcctggcccagatcggtgaccaatacgccgaccttttccttgctgctaagaatctttc
tgacgccatcctgctgtctgacattctccgcgtgaacactgaaatcaccaaggcccctctttcagcttcaatgattaagcggtatgatgagc
accaccaggacctgaccctgcttaaggcactcgtccggcagcagcttccggagaagtacaaggaaatcttctttgaccagtcaaagaa
tggatacgccggctacatcgacggaggtgcctcccaagaggaattttataagtttatcaaacctatccttgagaagatggacggcaccg
aagagctcctcgtgaaactgaatcgggaggatctgctgcggaagcagcgcactttcgacaatgggagcattccccaccagatccatctt
ggggagcttcacgccatccttcggcgccaagaggacttctacccctttcttaaggacaacagggagaagattgagaaaattctcactttc
cgcatcccctactacgtgggacccctcgccagaggaaatagccggtttgcttggatgaccagaaagtcagaagaaactatcactccct
ggaacttcgaagaggtggtggacaagggagccagcgctcagtcattcatcgaacggatgactaacttcgataagaacctccccaatg
agaaggtcctgccgaaacattccctgctctacgagtactttaccgtgtacaacgagctgaccaaggtgaaatatgtcaccgaagggatg
aggaagcccgcattcctgtcaggcgaacaaaagaaggcaattgtggaccttctgttcaagaccaatagaaaggtgaccgtgaagcag
ctgaaggaggactatttcaagaaaattgaatgcttcgactctgtggagattagcggggtcgaagatcggttcaacgcaagcctgggtac

ctaccatgatctgcttaagatcatcaaggacaaggattttctggacaatgaggagaacgaggacatccttgaggacattgtcctgactctc
actctgttcgaggaccgggaaatgatcgaggagaggcttaagacctacgcccatctgttcgacgataaagtgatgaagcaacttaaac
ggagaagatataccggatggggacgccttagccgcaaactcatcaacggaatccgggacaaacagagcggaaagaccattcttgat
ttccttaaaagcgacggattcgctaatcgcaacttcatgcaacttatccatgatgattccctgacctttaaggaggacatccagaaggccc
aagtgtctggacaaggtgactcactgcacgagcatatcgcaaatctggctggttcacccgctattaagaagggtattctccagaccgtga
aagtcgtggacgagctggtcaaggtgatgggtcgccataaaccagagaacattgtcatcgagatggccagggaaaaccagactacc
cagaagggacagaagaacagcagggagcggatgaaaagaattgaggaagggattaaggagctcgggtcacagatccttaaaga
gcacccggtggaaaacacccagcttcagaatgagaagctctatctgtactaccttcaaaatggacgcgatatgtatgtggaccaagag
cttgatatcaacaggctctcagactacgacgtggacgccatcgtccctcagagcttcctcaaagacgactcaattgacaataaggtgctg
actcgctcagacaagaaccggggaaagtcagataacgtgccctcagaggaagtcgtgaaaaagatgaagaactattggcgccagct
tctgaacgcaaagctgatcactcagcggaagttcgacaatctcactaaggctgagaggggcggactgagcgaactggacaaagcag
gattcattaaacggcaacttgtggagactcggcagattactaaacatgtcgcccaaatccttgactcacgcatgaataccaagtacgacg
aaaacgacaaacttatccgcgaggtgaaggtgattaccctgaagtccaagctggtcagcgatttcagaaaggactttcaattctacaaa
gtgcgggagatcaataactatcatcatgctcatgacgcatatctgaatgccgtggtgggaaccgccctgatcaagaagtacccaaagct
ggaaagcgagttcgtgtacggagactacaaggtctacgacgtgcgcaagatgattgccaaatctgagcaggagatcggaaaggcca
ccgcaaagtacttcttctacagcaacatcatgaatttcttcaagaccgaaatcacccttgcaaacggtgagatccggaagaggccgctc
atcgagactaatggggagactggcgaaatcgtgtgggacaagggcagagatttcgctaccgtgcgcaaagtgctttctatgcctcaagt
gaacatcgtgaagaaaaccgaggtgcaaaccggaggcttttctaaggaatcaatcctccccaagcgcaactccgacaagctcattgc
aaggaagaaggattgggaccctaagaagtacggcggattcgattcaccaactgtggcttattctgtcctggtcgtggctaaggtggaaa
aaggaaagtctaagaagctcaagagcgtgaaggaactgctgggtatcaccattatggagcgcagctccttcgagaagaacccaattg
actttctcgaagccaaaggttacaaggaagtcaagaaggaccttatcatcaagctcccaaagtatagcctgttcgaactggagaatggg
cggaagcggatgctcgcctccgctggcgaacttcagaagggtaatgagctggctctcccctccaagtacgtgaatttcctctaccttgcaa
gccattacgagaagctgaaggggagccccgaggacaacgagcaaaagcaactgtttgtggagcagcataagcattatctggacga
gatcattgagcagatttccgagttttctaaacgcgtcattctcgctgatgccaacctcgataaagtccttagcgcatacaataagcacagag
acaaaccaattcgggagcaggctgagaatatcatccacctgttcaccctcaccaatcttggtgcccctgccgcattcaagtacttcgaca
ccaccatcgaccggaaacgctatacctccaccaaagaagtgctggacgccaccctcatccaccagagcatcaccggactttacgaaa
ctcggattgacctctcacagctcggaggggatggtggcggaggctcgccaaaaaagaagagaaaggtagacccaaagaaaaaac
gaaaagtagatccgaaaaagaagaggaaggtgggatccctgcccacctgcagctgtcttgatcgagttatacaaaaagacaaaggc
ccatattatacacaccttggggcaggaccaagtgttgctgctgtcagggaaatcatggagaataggtatggtcaaaaaggaaacgcaa
taaggatagaaatagtagtgtacaccggtaaagaagggaaaagctctcatgggtgtccaattgctaagtgggttttaagaagaagcagt
gatgaagaaaaagttctttgtttggtccggcagcgtacaggccaccactgtccaactgctgtgatggtggtgctcatcatggtgtgggatgg
catccctcttccaatggccgaccggctatacacagagctcacagagaatctaaagtcatacaatgggcaccctaccgacagaagatgc
accctcaatgaaaatcgtacctgtacatgtcaaggaattgatccagagacttgtggagcttcattctcttttggctgttcatggagtatgtacttt
aatggctgtaagtttggtagaagcccaagccccagaagatttagaattgatccaagctctcccttacatgaaaaaaaccttgaagataac
ttacagagtttggctacacgattagctccaatttataagcagtatgctccagtagcttaccaaaatcaggtggaatatgaaaatgttgcccg
agaatgtcggcttggcagcaaggaaggtcgacccttctctggggtcactgcttgcctggacttctgtgctcatccccacagggacattcac
aacatgaataatggaagcactgtggtttgtaccttaactcgagaagataaccgctctttgggtgttattcctcaagatgagcagctccatgtg
ctacctctttataagctttcagacacagatgagtttggctccaaggaaggaatggaagccaagatcaaatctggggccatcgaggtcctg
gcaccccgccgcaaaaaaagaacgtgtttcactcagcctgttccccgttctggaaagaagagggctgcgatgatgacagaggttcttgc
acataagataagggcagtggaaaagaaacctattccccgaatcaagcggaagaataactcaacaacaacaaacaacagtaagcct
tcgtcactgccaaccttagggagtaacactgagaccgtgcaacctgaagtaaaaagtgaaaccgaaccccattttatcttaaaaagttc
agacaacactaaaacttattcgctgatgccatccgctcctcacccagtgaaagaggcatctccaggcttctcctggtccccgaagactgc
ttcagccacaccagctccactgaagaatgacgcaacagcctcatgcgggttttcagaaagaagcagcactccccactgtacgatgcctt
cgggaagactcagtggtgccaatgctgcagctgctgatggccctggcatttcacagcttggcgaagtggctcctctccccaccctgtctgc
tcctgtgatggagcccctcattaattctgagccttccactggtgtgactgagccgctaacgcctcatcagccaaaccaccagccctccttcc
tcacctctcctcaagaccttgcctcttctccaatggaagaagatgagcagcattctgaagcagatgagcctccatcagacgaacccctat
ctgatgacccccctgtcacctgctgaggagaaattgccccacattgatgagtattggtcagacagtgagcacatctttttggatgcaaatatt
ggtggggtggccatcgcacctgctcacggctcggttttgattgagtgtgcccggcgagagctgcacgctaccactcctgttgagcacccc
aaccgtaatcatccaacccgcctctcccttgtcttttaccagcacaaaaacctaaataagccccaacatggttttgaactaaacaagatta
agtttgaggctaaagaagctaagaataagaaaatgaaggcctcagagcaaaaagaccaggcagctaatgaaggtccagaacagtc
ctctgaagtaaatgaattgaaccaaattccttctcataaagcattaacattaacccatgacaatgttgtcaccgtgtccccttatgctctcaca
cacgttgcggggccctataaccattgggtcgagggcagaggaagtctgctaacatgcggtgacgtcgaggagaatcctggcccagtg
agcaagggcgaggagctgttcaccggggtggtgcccatcctggtcgagctggacggcgacgtaaacggccacaagttcagcgtgtcc
ggcgagggcgagggcgatgccacctacggcaagctgaccctgaagttcatctgcaccaccggcaagctgcccgtgccctggcccac
cctcgtgaccaccctgacctacggcgtgcagtgcttcagccgctaccccgaccacatgaagcagcacgacttcttcaagtccgccatgc
ccgaaggctacgtccaggagcgcaccatcttcttcaaggacgacggcaactacaagacccgcgccgaggtgaagttcgagggcga

caccctggtgaaccgcatcgagctgaagggcatcgacttcaaggaggacggcaacatcctggggcacaagctggagtacaactaca
acagccacaacgtctatatcatggccgacaagcagaagaacggcatcaaggtgaacttcaagatccgccacaacatcgaggacgg
cagcgtgcagctcgccgaccactaccagcagaacacccccatcggcgacggccccgtgctgctgcccgacaaccactacctgagca
cccagtccgccctgagcaaagaccccaacgagaagcgcgatcacatggtcctgctggagttcgtgaccgccgccgggatcactctcg
gcatggacgagctgtacaaggaattcgatatcaagcttatcgataatcaacctctggattacaaaatttgtgaaagattgactggtattctta
actatgttgctccttttacgctatgtggatacgctgctttaatgcctttgtatcatgctattgcttcccgtatggctttcattttctcctccttgtataaat
cctggttgctgtctctttatgaggagttgtggcccgttgtcaggcaacgtggcgtggtgtgcactgtgtttgctgacgcaacccccactggttg
gggcattgccaccacctgtcagctcctttccgggactttcgctttccccctccctattgccacggcggaactcatcgccgcctgccttgcccg
ctgctggacaggggctcggctgttgggcactgacaattccgtggtgttgtcggggaaatcatcgtcctttccttggctgctcgcctgtgttgcc
acctggattctgcgcgggacgtccttctgctacgtcccttcggccctcaatccagcggaccttccttcccgcggcctgctgccggctctgcg
gcctcttccgcgtcttcgccttcgccctcagacgagtcggatctccctttgggccgcctccccgcatcgataccgtcgacctcgagacctag
aaaaacatggagcaatcacaagtagcaatacagcagctaccaatgctgattgtgcctggctagaagcacaagaggaggaggaggt
gggttttccagtcacacctcaggtacctttaagaccaatgacttacaaggcagctgtagatcttagccactttttaaaagaaaaggggggga
ctggaagggctaattcactcccaacgaagacaagatatccttgatctgtggatctaccacacacaaggctacttccctgattggcagaac
tacacaccagggccagggatcagatatccactgacctttggatggtgctacaagctagtaccagttgagcaagagaaggtagaagaa
gccaatgaaggagagaacacccgcttgttacaccctgtgagcctgcatgggatggatgacccggagagagaagtattagagtggag
gtttgacagccgcctagcatttcatcacatggcccgagagctgcatccggactgtactgggtctctctggttagaccagatctgagcctgg
gagctctctggctaactagggaacccactgcttaagcctcaataaagcttgccttgagtgcttcaagtagtgtgtgcccgtctgttgtgtgact
ctggtaactagagatccctcagacccttttagtcagtgtggaaaatctctagcagggcccgtttaaacccgctgatcagcctcgactgtgc
cttctagttgccagccatctgttgtttgcccctcccccgtgccttccttgaccctggaaggtgccactcccactgtcctttcctaataaaatgag
gaaattgcatcgcattgtctgagtaggtgtcattctattctggggggtggggtggggcaggacagcaagggggaggattgggaagaca
atagcaggcatgctggggatgcggtgggctctatggcttctgaggcggaaagaaccagctggggctctagggggtatccccacgcgc
cctgtagcggcgcattaagcgcggcgggtgtggtggttacgcgcagcgtgaccgctacacttgccagcgccctagcgcccgctccttc
gctttcttcccttcctttctcgccacgttcgccggctttccccgtcaagctctaaatcgggggctccctttagggttccgatttagtgctttacggc
acctcgaccccaaaaaacttgattagggtgatggttcacgtagtgggccatcgccctgatagacggtttttcgccctttgacgttggagtcc
acgttctttaatagtggactcttgttccaaactggaacaacactcaaccctatctcggtctattcttttgatttataagggattttgccgatttcgg
cctattggttaaaaaatgagctgatttaacaaaaatttaacgcgaattaattctgtggaatgtgtgtcagttagggtgtggaaagtccccag
gctccccagcaggcagaagtatgcaaagcatgcatctcaattagtcagcaaccatagtcccgcccctaactccgcccatcccgcccct
aactccgcccagttccgcccattctccgccccatggctgactaattttttttatttatgcagaggccgaggccgcctctgcctctgagctattcc
agaagtagtgaggaggcttttttggaggcctaggcttttgcaaaaagctcccgggagcttgtatatccattttcggatctgatcagcacgtgt
tgacaattaatcatcggcatagtatatcggcatagtataatacgacaaggtgaggaactaaaccatggccaagttgaccagtgccgttcc
ggtgctcaccgcgcgcgacgtcgccggagcggtcgagttctggaccgaccggctcgggttctcccgggacttcgtggaggacgacttc
gccggtgtggtccgggacgacgtgaccctgttcatcagcgcggtccaggaccaggtggtgccggacaacaccctggcctgggtgtgg
gtgcgcggcctggacgagctgtacgccgagtggtcggaggtcgtgtccacgaacttccgggacgcctccgggccggccatgaccgag
atcggcgagcagccgtgggggcgggagttcgccctgcgcgacccggccggcaactgcgtgcacttcgtggccgaggagcaggact
gacacgtgctacgagatttcgattccaccgccgccttctatgaaaggttgggcttcggaatcgttttccgggacgccggctggatgatcctc
cagcgcggggatctcatgctggagttcttcgcccaccccaacttgtttattgcagcttataatggttacaaataaagcaatagcatcacaaa
tttcacaaataaagcatttttttcactgcattctagttgtggtttgtccaaactcatcaatgtatcttatcatgtctgtataccgtcgacctctagcta
gagcttggcgtaatcatggtcatagctgtttcctgtgtgaaattgttatccgctcacaattccacacaacatacgagccggaagcataaagt
gtaaagcc

**P3-Lenti-dCas9-Tet1IM-GFP (Addgene #190730)**

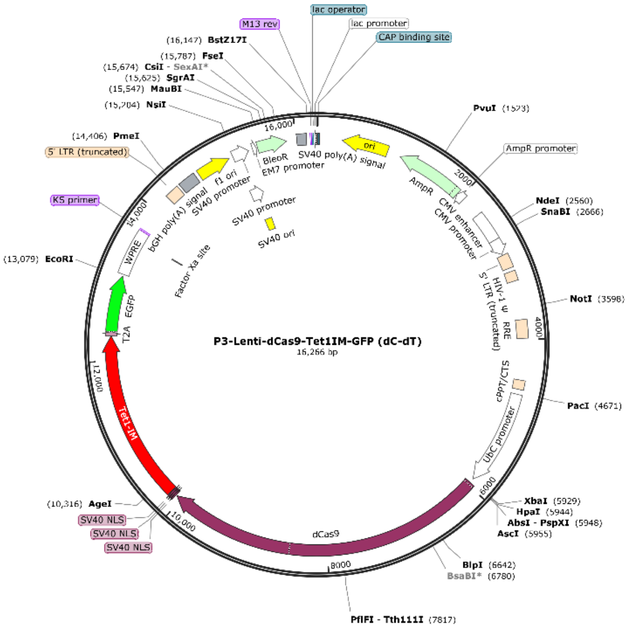


tggggtgcctaatgagtgagctaactcacattaattgcgttgcgctcactgcccgctttccagtcgggaaacctgtcgtgccagctgcatta
atgaatcggccaacgcgcggggagaggcggtttgcgtattgggcgctcttccgcttcctcgctcactgactcgctgcgctcggtcgttcgg
ctgcggcgagcggtatcagctcactcaaaggcggtaatacggttatccacagaatcaggggataacgcaggaaagaacatgtgagc
aaaaggccagcaaaaggccaggaaccgtaaaaaggccgcgttgctggcgtttttccataggctccgcccccctgacgagcatcaca
aaaatcgacgctcaagtcagaggtggcgaaacccgacaggactataaagataccaggcgtttccccctggaagctccctcgtgcgct
ctcctgttccgaccctgccgcttaccggatacctgtccgcctttctcccttcgggaagcgtggcgctttctcatagctcacgctgtaggtatctc
agttcggtgtaggtcgttcgctccaagctgggctgtgtgcacgaaccccccgttcagcccgaccgctgcgccttatccggtaactatcgtct
tgagtccaacccggtaagacacgacttatcgccactggcagcagccactggtaacaggattagcagagcgaggtatgtaggcggtgc
tacagagttcttgaagtggtggcctaactacggctacactagaagaacagtatttggtatctgcgctctgctgaagccagttaccttcggaa
aaagagttggtagctcttgatccggcaaacaaaccaccgctggtagcggtggtttttttgtttgcaagcagcagattacgcgcagaaaaa
aaggatctcaagaagatcctttgatcttttctacggggtctgacgctcagtggaacgaaaactcacgttaagggattttggtcatgagattat
caaaaaggatcttcacctagatccttttaaattaaaaatgaagttttaaatcaatctaaagtatatatgagtaaacttggtctgacagttacca
atgcttaatcagtgaggcacctatctcagcgatctgtctatttcgttcatccatagttgcctgactccccgtcgtgtagataactacgatacgg
gagggcttaccatctggccccagtgctgcaatgataccgcgagacccacgctcaccggctccagatttatcagcaataaaccagccag
ccggaagggccgagcgcagaagtggtcctgcaactttatccgcctccatccagtctattaattgttgccgggaagctagagtaagtagttc
gccagttaatagtttgcgcaacgttgttgccattgctacaggcatcgtggtgtcacgctcgtcgtttggtatggcttcattcagctccggttccc
aacgatcaaggcgagttacatgatcccccatgttgtgcaaaaaagcggttagctccttcggtcctccgatcgttgtcagaagtaagttggc
cgcagtgttatcactcatggttatggcagcactgcataattctcttactgtcatgccatccgtaagatgcttttctgtgactggtgagtactcaac
caagtcattctgagaatagtgtatgcggcgaccgagttgctcttgcccggcgtcaatacgggataataccgcgccacatagcagaacttt
aaaagtgctcatcattggaaaacgttcttcggggcgaaaactctcaaggatcttaccgctgttgagatccagttcgatgtaacccactcgt
gcacccaactgatcttcagcatcttttactttcaccagcgtttctgggtgagcaaaaacaggaaggcaaaatgccgcaaaaaagggaat
aagggcgacacggaaatgttgaatactcatactcttcctttttcaatattattgaagcatttatcagggttattgtctcatgagcggatacatatt
tgaatgtatttagaaaaataaacaaataggggttccgcgcacatttccccgaaaagtgccacctgacgtcgacggatcgggagatctcc
cgatcccctatggtgcactctcagtacaatctgctctgatgccgcatagttaagccagtatctgctccctgcttgtgtgttggaggtcgctgag
tagtgcgcgagcaaaatttaagctacaacaaggcaaggcttgaccgacaattgcatgaagaatctgcttagggttaggcgttttgcgctg
cttcgcgatgtacgggccagatatacgcgttgacattgattattgactagttattaatagtaatcaattacggggtcattagttcatagcccat
atatggagttccgcgttacataacttacggtaaatggcccgcctggctgaccgcccaacgacccccgcccattgacgtcaataatgacgt
atgttcccatagtaacgccaatagggactttccattgacgtcaatgggtggagtatttacggtaaactgcccacttggcagtacatcaagtg
tatcatatgccaagtacgccccctattgacgtcaatgacggtaaatggcccgcctggcattatgcccagtacatgaccttatgggactttcc
tacttggcagtacatctacgtattagtcatcgctattaccatggtgatgcggttttggcagtacatcaatgggcgtggatagcggtttgactca
cggggatttccaagtctccaccccattgacgtcaatgggagtttgttttggcaccaaaatcaacgggactttccaaaatgtcgtaacaactc
cgccccattgacgcaaatgggcggtaggcgtgtacggtgggaggtctatataagcagcgcgttttgcctgtactgggtctctctggttaga
ccagatctgagcctgggagctctctggctaactagggaacccactgcttaagcctcaataaagcttgccttgagtgcttcaagtagtgtgtg
cccgtctgttgtgtgactctggtaactagagatccctcagacccttttagtcagtgtggaaaatctctagcagtggcgcccgaacagggact

tgaaagcgaaagggaaaccagaggagctctctcgacgcaggactcggcttgctgaagcgcgcacggcaagaggcgaggggcgg
cgactggtgagtacgccaaaaattttgactagcggaggctagaaggagagagatgggtgcgagagcgtcagtattaagcgggggag
aattagatcgcgatgggaaaaaattcggttaaggccagggggaaagaaaaaatataaattaaaacatatagtatgggcaagcaggg
agctagaacgattcgcagttaatcctggcctgttagaaacatcagaaggctgtagacaaatactgggacagctacaaccatcccttcag
acaggatcagaagaacttagatcattatataatacagtagcaaccctctattgtgtgcatcaaaggatagagataaaagacaccaagg
aagctttagacaagatagaggaagagcaaaacaaaagtaagaccaccgcacagcaagcggccgctgatcttcagacctggagga
ggagatatgagggacaattggagaagtgaattatataaatataaagtagtaaaaattgaaccattaggagtagcacccaccaaggca
aagagaagagtggtgcagagagaaaaaagagcagtgggaataggagctttgttccttgggttcttgggagcagcaggaagcactatg
ggcgcagcgtcaatgacgctgacggtacaggccagacaattattgtctggtatagtgcagcagcagaacaatttgctgagggctattga
ggcgcaacagcatctgttgcaactcacagtctggggcatcaagcagctccaggcaagaatcctggctgtggaaagatacctaaagga
tcaacagctcctggggatttggggttgctctggaaaactcatttgcaccactgctgtgccttggaatgctagttggagtaataaatctctgga
acagatttggaatcacacgacctggatggagtgggacagagaaattaacaattacacaagcttaatacactccttaattgaagaatcgc
aaaaccagcaagaaaagaatgaacaagaattattggaattagataaatgggcaagtttgtggaattggtttaacataacaaattggctg
tggtatataaaattattcataatgatagtaggaggcttggtaggtttaagaatagtttttgctgtactttctatagtgaatagagttaggcaggg
atattcaccattatcgtttcagacccacctcccaaccccgaggggacccgacaggcccgaaggaatagaagaagaaggtggagaga
gagacagagacagatccattcgattagtgaacggatcggcactgcgtgcgccaattctgcagacaaatggcagtattcatccacaatttt
aaaagaaaaggggggattggggggtacagtgcaggggaaagaatagtagacataatagcaacagacatacaaactaaagaatta
caaaaacaaattacaaaaattcaaaattttcgggtttattacagggacagcagagatccagtttggttaattaacccgtgtcggctccaga
tctggcctccgcgccgggttttggcgcctcccgcgggcgcccccctcctcacggcgagcgctgccacgtcagacgaagggcgcagcg
agcgtcctgatccttccgcccggacgctcaggacagcggcccgctgctcataagactcggccttagaaccccagtatcagcagaagg
acattttaggacgggacttgggtgactctagggcactggttttctttccagagagcggaacaggcgaggaaaagtagtcccttctcggcg
attctgcggagggatctccgtggggcggtgaacgccgatgattatataaggacgcgccgggtgtggcacagctagttccgtcgcagcc
gggatttgggtcgcggttcttgtttgtggatcgctgtgatcgtcacttggtgagtagcgggctgctgggctggccggggctttcgtggccgcc
gggccgctcggtgggacggaagcgtgtggagagaccgccaagggctgtagtctgggtccgcgagcaaggttgccctgaactggggg
ttggggggagcgcagcaaaatggcggctgttcccgagtcttgaatggaagacgcttgtgaggcgggctgtgaggtcgttgaaacaagg
tgggggcatggtgggcggcaagaacccaaggtcttgaggccttcgctaatgcgggaaagctcttattcgggtgagatgggctggggc
accatctggggaccctgacgtgaagtttgtcactgactggagaactcggtttgtcgtctgttgcgggggcggcagttatggcggtgccgttg
ggcagtgcacccgtacctttgggagcgcgcgccctcgtcgtgtcgtgacgtcacccgttctgttggcttataatgcagggtggggccacct
gccggtaggtgtgcggtaggcttttctccgtcgcaggacgcagggttcgggcctagggtaggctctcctgaatcgacaggcgccggacc
tctggtgaggggagggataagtgaggcgtcagtttctttggtcggttttatgtacctatcttcttaagtagctgaagctccggttttgaactatgc
gctcggggttggcgagtgtgttttgtgaagttttttaggcaccttttgaaatgtaatcatttgggtcaatatgtaattttcagtgttagactagtaaa
ttgtccgctaaattctggccgtttttggcttttttgttagacgaagcttgggctgcaggtcgactctagaggatccagttaacctcgagggcgc
gcccatggacaagaagtattctatcggactggccatcgggactaatagcgtcgggtgggccgtgatcactgacgagtacaaggtgccc
tctaagaagttcaaggtgctcgggaacaccgaccggcattccatcaagaaaaatctgatcggagctctcctctttgattcaggggagacc
gctgaagcaacccgcctcaagcggactgctagacggcggtacaccaggaggaagaaccggatttgttaccttcaagagatattctcca
acgaaatggcaaaggtcgacgacagcttcttccataggctggaagaatcattcctcgtggaagaggataagaagcatgaacggcatc
ccatcttcggtaatatcgtcgacgaggtggcctatcacgagaaatacccaaccatctaccatcttcgcaaaaagctggtggactcaacc
gacaaggcagacctccggcttatctacctggccctggcccacatgatcaagttcagaggccacttcctgatcgagggcgacctcaatcc
tgacaatagcgatgtggataaactgttcatccagctggtgcagacttacaaccagctctttgaagagaaccccatcaatgcaagcggag
tcgatgccaaggccattctgtcagcccggctgtcaaagagccgcagacttgagaatcttatcgctcagctgccgggtgaaaagaaaaat
ggactgttcgggaacctgattgctctttcacttggcctgactcccaatttcaagtctaatttcgacctggcagaggatgccaagctgcaactg
tccaaggacacctatgatgacgatctcgacaacctcctggcccagatcggtgaccaatacgccgaccttttccttgctgctaagaatctttc
tgacgccatcctgctgtctgacattctccgcgtgaacactgaaatcaccaaggcccctctttcagcttcaatgattaagcggtatgatgagc
accaccaggacctgaccctgcttaaggcactcgtccggcagcagcttccggagaagtacaaggaaatcttctttgaccagtcaaagaa
tggatacgccggctacatcgacggaggtgcctcccaagaggaattttataagtttatcaaacctatccttgagaagatggacggcaccg
aagagctcctcgtgaaactgaatcgggaggatctgctgcggaagcagcgcactttcgacaatgggagcattccccaccagatccatctt
ggggagcttcacgccatccttcggcgccaagaggacttctaccccttcttaaggacaacagggagaagattgagaaaattctcactttc
cgcatcccctactacgtgggacccctcgccagaggaaatagccggtttgcttggatgaccagaaagtcagaagaaactatcactccct
ggaacttcgaagaggtggtggacaagggagccagcgctcagtcattcatcgaacggatgactaacttcgataagaacctccccaatg
agaaggtcctgccgaaacattccctgctctacgagtactttaccgtgtacaacgagctgaccaaggtgaaatatgtcaccgaagggatg
aggaagcccgcattcctgtcaggcgaacaaaagaaggcaattgtggaccttctgttcaagaccaatagaaaggtgaccgtgaagcag
ctgaaggaggactatttcaagaaaattgaatgcttcgactctgtggagattagcggggtcgaagatcggttcaacgcaagcctgggtac
ctaccatgatctgcttaagatcatcaaggacaaggattttctggacaatgaggagaacgaggacatccttgaggacattgtcctgactctc
actctgttcgaggaccgggaaatgatcgaggagaggcttaagacctacgcccatctgttcgacgataaagtgatgaagcaacttaaac
ggagaagatataccggatggggacgccttagccgcaaactcatcaacggaatccgggacaaacagagcggaaagaccattcttgat
ttccttaaaagcgacggattcgctaatcgcaacttcatgcaacttatccatgatgattccctgacctttaaggaggacatccagaaggccc

aagtgtctggacaaggtgactcactgcacgagcatatcgcaaatctggctggttcacccgctattaagaagggtattctccagaccgtga
aagtcgtggacgagctggtcaaggtgatgggtcgccataaaccagagaacattgtcatcgagatggccagggaaaaccagactacc
cagaagggacagaagaacagcagggagcggatgaaaagaattgaggaagggattaaggagctcgggtcacagatccttaaaga
gcacccggtggaaaacacccagcttcagaatgagaagctctatctgtactaccttcaaaatggacgcgatatgtatgtggaccaagag
cttgatatcaacaggctctcagactacgacgtggacgccatcgtccctcagagcttcctcaaagacgactcaattgacaataaggtgctg
actcgctcagacaagaaccggggaaagtcagataacgtgccctcagaggaagtcgtgaaaaagatgaagaactattggcgccagct
tctgaacgcaaagctgatcactcagcggaagttcgacaatctcactaaggctgagaggggcggactgagcgaactggacaaagcag
gattcattaaacggcaacttgtggagactcggcagattactaaacatgtcgcccaaatccttgactcacgcatgaataccaagtacgacg
aaaacgacaaacttatccgcgaggtgaaggtgattaccctgaagtccaagctggtcagcgatttcagaaaggactttcaattctacaaa
gtgcgggagatcaataactatcatcatgctcatgacgcatatctgaatgccgtggtgggaaccgccctgatcaagaagtacccaaagct
ggaaagcgagttcgtgtacggagactacaaggtctacgacgtgcgcaagatgattgccaaatctgagcaggagatcggaaaggcca
ccgcaaagtacttcttctacagcaacatcatgaatttcttcaagaccgaaatcacccttgcaaacggtgagatccggaagaggccgctc
atcgagactaatggggagactggcgaaatcgtgtgggacaagggcagagatttcgctaccgtgcgcaaagtgctttctatgcctcaagt
gaacatcgtgaagaaaaccgaggtgcaaaccggaggcttttctaaggaatcaatcctccccaagcgcaactccgacaagctcattgc
aaggaagaaggattgggaccctaagaagtacggcggattcgattcaccaactgtggcttattctgtcctggtcgtggctaaggtggaaa
aaggaaagtctaagaagctcaagagcgtgaaggaactgctgggtatcaccattatggagcgcagctccttcgagaagaacccaattg
actttctcgaagccaaaggttacaaggaagtcaagaaggaccttatcatcaagctcccaaagtatagcctgttcgaactggagaatggg
cggaagcggatgctcgcctccgctggcgaacttcagaagggtaatgagctggctctcccctccaagtacgtgaatttcctctaccttgcaa
gccattacgagaagctgaaggggagccccgaggacaacgagcaaaagcaactgtttgtggagcagcataagcattatctggacga
gatcattgagcagatttccgagttttctaaacgcgtcattctcgctgatgccaacctcgataaagtccttagcgcatacaataagcacagag
acaaaccaattcgggagcaggctgagaatatcatccacctgttcaccctcaccaatcttggtgcccctgccgcattcaagtacttcgaca
ccaccatcgaccggaaacgctatacctccaccaaagaagtgctggacgccaccctcatccaccagagcatcaccggactttacgaaa
ctcggattgacctctcacagctcggaggggatggtggcggaggctcgccaaaaaagaagagaaaggtagacccaaagaaaaaac
gaaaagtagatccgaaaaagaagaggaaggtgggatccctgcccacctgcagctgtcttgatcgagttatacaaaaagacaaaggc
ccatattatacacaccttggggcaggaccaagtgttgctgctgtcagggaaatcatggagaataggtatggtcaaaaaggaaacgcaa
taaggatagaaatagtagtgtacaccggtaaagaagggaaaagctctcatgggtgtccaattgctaagtgggttttaagaagaagcagt
gatgaagaaaaagttctttgtttggtccggcagcgtacaggccaccactgtccaactgctgtgatggtggtgctcatcatggtgtgggatgg
catccctcttccaatggccgaccggctatacacagagctcacagagaatctaaagtcatacaatgggcaccctaccgacagaagatgc
accctcaatgaaaatcgtacctgtacatgtcaaggaattgatccagagacttgtggagcttcattctcttttggctgttcatggagtatgtacttt
aatggctgtaagtttggtagaagcccaagccccagaagatttagaattgatccaagctctcccttacatgaaaaaaaccttgaagataac
ttacagagtttggctacacgattagctccaatttataagcagtatgctccagtagcttaccaaaatcaggtggaatatgaaaatgttgcccg
agaatgtcggcttggcagcaaggaaggtcgacccttctctggggtcactgcttgcctggacttctgtgctcatccctacagggccattcac
aacatgaataatggaagcactgtggtttgtaccttaactcgagaagataaccgctctttgggtgttattcctcaagatgagcagctccatgtg
ctacctctttataagctttcagacacagatgagtttggctccaaggaaggaatggaagccaagatcaaatctggggccatcgaggtcctg
gcaccccgccgcaaaaaaagaacgtgtttcactcagcctgttccccgttctggaaagaagagggctgcgatgatgacagaggttcttgc
acataagataagggcagtggaaaagaaacctattccccgaatcaagcggaagaataactcaacaacaacaaacaacagtaagcct
tcgtcactgccaaccttagggagtaacactgagaccgtgcaacctgaagtaaaaagtgaaaccgaaccccattttatcttaaaaagttc
agacaacactaaaacttattcgctgatgccatccgctcctcacccagtgaaagaggcatctccaggcttctcctggtccccgaagactgc
ttcagccacaccagctccactgaagaatgacgcaacagcctcatgcgggttttcagaaagaagcagcactccccactgtacgatgcctt
cgggaagactcagtggtgccaatgctgcagctgctgatggccctggcatttcacagcttggcgaagtggctcctctccccaccctgtctgc
tcctgtgatggagcccctcattaattctgagccttccactggtgtgactgagccgctaacgcctcatcagccaaaccaccagccctccttcc
tcacctctcctcaagaccttgcctcttctccaatggaagaagatgagcagcattctgaagcagatgagcctccatcagacgaacccctat
ctgatgacccccctgtcacctgctgaggagaaattgccccacattgatgagtattggtcagacagtgagcacatctttttggatgcaaatatt
ggtggggtggccatcgcacctgctcacggctcggttttgattgagtgtgcccggcgagagctgcacgctaccactcctgttgagcacccc
aaccgtaatcatccaacccgcctctcccttgtcttttaccagcacaaaaacctaaataagccccaacatggttttgaactaaacaagatta
agtttgaggctaaagaagctaagaataagaaaatgaaggcctcagagcaaaaagaccaggcagctaatgaaggtccagaacagtc
ctctgaagtaaatgaattgaaccaaattccttctcataaagcattaacattaacccatgacaatgttgtcaccgtgtccccttatgctctcaca
cacgttgcggggccctataaccattgggtcgagggcagaggaagtctgctaacatgcggtgacgtcgaggagaatcctggcccagtg
agcaagggcgaggagctgttcaccggggtggtgcccatcctggtcgagctggacggcgacgtaaacggccacaagttcagcgtgtcc
ggcgagggcgagggcgatgccacctacggcaagctgaccctgaagttcatctgcaccaccggcaagctgcccgtgccctggcccac
cctcgtgaccaccctgacctacggcgtgcagtgcttcagccgctaccccgaccacatgaagcagcacgacttcttcaagtccgccatgc
ccgaaggctacgtccaggagcgcaccatcttcttcaaggacgacggcaactacaagacccgcgccgaggtgaagttcgagggcga
caccctggtgaaccgcatcgagctgaagggcatcgacttcaaggaggacggcaacatcctggggcacaagctggagtacaactaca
acagccacaacgtctatatcatggccgacaagcagaagaacggcatcaaggtgaacttcaagatccgccacaacatcgaggacgg
cagcgtgcagctcgccgaccactaccagcagaacacccccatcggcgacggccccgtgctgctgcccgacaaccactacctgagca
cccagtccgccctgagcaaagaccccaacgagaagcgcgatcacatggtcctgctggagttcgtgaccgccgccgggatcactctcg

gcatggacgagctgtacaaggaattcgatatcaagcttatcgataatcaacctctggattacaaaatttgtgaaagattgactggtattctta
actatgttgctccttttacgctatgtggatacgctgctttaatgcctttgtatcatgctattgcttcccgtatggctttcattttctcctccttgtataaat
cctggttgctgtctctttatgaggagttgtggcccgttgtcaggcaacgtggcgtggtgtgcactgtgtttgctgacgcaacccccactggttg
gggcattgccaccacctgtcagctcctttccgggactttcgctttccccctccctattgccacggcggaactcatcgccgcctgccttgcccg
ctgctggacaggggctcggctgttgggcactgacaattccgtggtgttgtcggggaaatcatcgtcctttccttggctgctcgcctgtgttgcc
acctggattctgcgcgggacgtccttctgctacgtcccttcggccctcaatccagcggaccttccttcccgcggcctgctgccggctctgcg
gcctcttccgcgtcttcgccttcgccctcagacgagtcggatctccctttgggccgcctccccgcatcgataccgtcgacctcgagacctag
aaaaacatggagcaatcacaagtagcaatacagcagctaccaatgctgattgtgcctggctagaagcacaagaggaggaggaggt
gggttttccagtcacacctcaggtacctttaagaccaatgacttacaaggcagctgtagatcttagccactttttaaaagaaaaggggga
ctggaagggctaattcactcccaacgaagacaagatatccttgatctgtggatctaccacacacaaggctacttccctgattggcagaac
tacacaccagggccagggatcagatatccactgacctttggatggtgctacaagctagtaccagttgagcaagagaaggtagaagaa
gccaatgaaggagagaacacccgcttgttacaccctgtgagcctgcatgggatggatgacccggagagagaagtattagagtggag
gtttgacagccgcctagcatttcatcacatggcccgagagctgcatccggactgtactgggtctctctggttagaccagatctgagcctgg
gagctctctggctaactagggaacccactgcttaagcctcaataaagcttgccttgagtgcttcaagtagtgtgtgcccgtctgttgtgtgact
ctggtaactagagatccctcagacccttttagtcagtgtggaaaatctctagcagggcccgtttaaacccgctgatcagcctcgactgtgc
cttctagttgccagccatctgttgtttgcccctcccccgtgccttccttgaccctggaaggtgccactcccactgtcctttcctaataaaatgag
gaaattgcatcgcattgtctgagtaggtgtcattctattctggggggtggggtggggcaggacagcaagggggaggattgggaagaca
atagcaggcatgctggggatgcggtgggctctatggcttctgaggcggaaagaaccagctggggctctagggggtatccccacgcgc
cctgtagcggcgcattaagcgcggcgggtgtggtggttacgcgcagcgtgaccgctacacttgccagcgccctagcgcccgctcctttc
gctttcttcccttcctttctcgccacgttcgccggctttccccgtcaagctctaaatcgggggctccctttagggttccgatttagtgctttacggc
acctcgaccccaaaaaacttgattagggtgatggttcacgtagtgggccatcgccctgatagacggtttttcgccctttgacgttggagtcc
acgttctttaatagtggactcttgttccaaactggaacaacactcaaccctatctcggtctattcttttgatttataagggattttgccgatttcgg
cctattggttaaaaaatgagctgatttaacaaaaatttaacgcgaattaattctgtggaatgtgtgtcagttagggtgtggaaagtccccag
gctccccagcaggcagaagtatgcaaagcatgcatctcaattagtcagcaaccatagtcccgcccctaactccgcccatcccgcccct
aactccgcccagttccgcccattctccgccccatggctgactaattttttttatttatgcagaggccgaggccgcctctgcctctgagctattcc
agaagtagtgaggaggcttttttggaggcctaggcttttgcaaaaagctcccgggagcttgtatatccattttcggatctgatcagcacgtgt
tgacaattaatcatcggcatagtatatcggcatagtataatacgacaaggtgaggaactaaaccatggccaagttgaccagtgccgttcc
ggtgctcaccgcgcgcgacgtcgccggagcggtcgagttctggaccgaccggctcgggttctcccgggacttcgtggaggacgacttc
gccggtgtggtccgggacgacgtgaccctgttcatcagcgcggtccaggaccaggtggtgccggacaacaccctggcctgggtgtgg
gtgcgcggcctggacgagctgtacgccgagtggtcggaggtcgtgtccacgaacttccgggacgcctccgggccggccatgaccgag
atcggcgagcagccgtgggggcgggagttcgccctgcgcgacccggccggcaactgcgtgcacttcgtggccgaggagcaggact
gacacgtgctacgagatttcgattccaccgccgccttctatgaaaggttgggcttcggaatcgttttccgggacgccggctggatgatcctc
cagcgcggggatctcatgctggagttcttcgcccaccccaacttgtttattgcagcttataatggttacaaataaagcaatagcatcacaaa
tttcacaaataaagcatttttttcactgcattctagttgtggtttgtccaaactcatcaatgtatcttatcatgtctgtataccgtcgacctctagcta
gagcttggcgtaatcatggtcatagctgtttcctgtgtgaaattgttatccgctcacaattccacacaacatacgagccggaagcataaagt
gtaaagcc

**P3-pKLV2-U6gRNA(BbsI)-PGKpuro2A-mCherry**

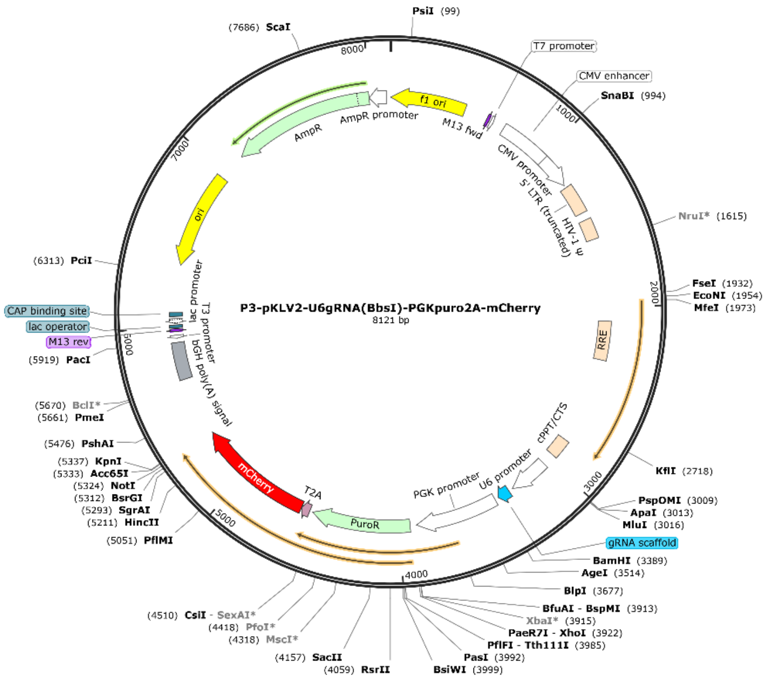


ctaaattgtaagcgttaatattttgttaaaattcgcgttaaatttttgttaaatcagctcattttttaaccaataggccgaaatcggcaaaatccct
tataaatcaaaagaatagaccgagatagggttgagtgttgttccagtttggaacaagagtccactattaaagaacgtggactccaacgtc
aaagggcgaaaaaccgtctatcagggcgatggcccactacgtgaaccatcaccctaatcaagttttttggggtcgaggtgccgtaaag
cactaaatcggaaccctaaagggagcccccgatttagagcttgacggggaaagccggcgaacgtggcgagaaaggaagggaag
aaagcgaaaggagcgggcgctagggcgctggcaagtgtagcggtcacgctgcgcgtaaccaccacacccgccgcgcttaatgcgc
cgctacagggcgcgtcccattcgccattcaggctgcgcaactgttgggaagggcgatcggtgcgggcctcttcgctattacgccagctgg
cgaaagggggatgtgctgcaaggcgattaagttgggtaacgccagggttttcccagtcacgacgttgtaaaacgacggccagtgagcg
cgcgtaatacgactcactatagggcgaattgactagttattaatagtaatcaattacggggtcattagttcatagcccatatatggagttccg
cgttacataacttacggtaaatggcccgcctggctgaccgcccaacgacccccgcccattgacgtcaataatgacgtatgttcccatagt
aacgccaatagggactttccattgacgtcaatgggtggagtatttacggtaaactgcccacttggcagtacatcaagtgtatcatatgcca
agtacgccccctattgacgtcaatgacggtaaatggcccgcctggcattatgcccagtacatgaccttatgggactttcctacttggcagta
catctacgtattagtcatcgctattaccatggtgatgcggttttggcagtacatcaatgggcgtggatagcggtttgactcacggggatttcca
agtctccaccccattgacgtcaatgggagtttgttttggcaccaaaatcaacgggactttccaaaatgtcgtaacaactccgccccattga
cgcaaatgggcggtaggcgtgtacggtgggaggtctatataagcagcgcgttttgcctgtactgggtctctctggttagaccagatctgag
cctgggagctctctggctaactagggaacccactgcttaagcctcaataaagcttgccttgagtgcttcaagtagtgtgtgcccgtctgttgt
gtgactctggtaactagagatccctcagacccttttagtcagtgtggaaaatctctagcagtggcgcccgaacagggacttgaaagcga
aagggaaaccagaggagctctctcgacgcaggactcggcttgctgaagcgcgcacggcaagaggcgaggggcggcgactggtga
gtacgccaaaaattttgactagcggaggctagaaggagagagatgggtgcgagagcgtcagtattaagcgggggagaattagatcg
cgatgggaaaaaattcggttaaggccagggggaaagaaaaaatataaattaaaacatatagtatgggcaagcagggagctagaac
gattcgcagttaatcctggcctgttagaaacatcagaaggctgtagacaaatactgggacagctacaaccatcccttcagacaggatca
gaagaacttagatcattatataatacagtagcaaccctctattgtgtgcatcaaaggatagagataaaagacaccaaggaagctttaga
caagatagaggaagagcaaaacaaaagtaagaccaccgcacagcaagcggccggccgcgctgatcttcagacctggaggagga
gatatgagggacaattggagaagtgaattatataaatataaagtagtaaaaattgaaccattaggagtagcacccaccaaggcaaag
agaagagtggtgcagagagaaaaaagagcagtgggaataggagctttgttccttgggttcttgggagcagcaggaagcactatgggc
gcagcgtcaatgacgctgacggtacaggccagacaattattgtctggtatagtgcagcagcagaacaatttgctgagggctattgaggc
gcaacagcatctgttgcaactcacagtctggggcatcaagcagctccaggcaagaatcctggctgtggaaagatacctaaaggatca
acagctcctggggatttggggttgctctggaaaactcatttgcaccactgctgtgccttggaatgctagttggagtaataaatctctggaaca
gatttggaatcacacgacctggatggagtgggacagagaaattaacaattacacaagcttaatacactccttaattgaagaatcgcaaa
accagcaagaaaagaatgaacaagaattattggaattagataaatgggcaagtttgtggaattggtttaacataacaaattggctgtggt
atataaaattattcataatgatagtaggaggcttggtaggtttaagaatagtttttgctgtactttctatagtgaatagagttaggcagggatatt
caccattatcgtttcagacccacctcccaaccccgaggggacccgacaggcccgaaggaatagaagaagaaggtggagagagag
acagagacagatccattcgattagtgaacggatcggcactgcgtgcgccaattctgcagacaaatggcagtattcatccacaattttaaa

agaaaagggggggattgggggggtacagtgcaggggaaagaatagtagacataatagcaacagacatacaaactaaagaattacaa
aaacaaattacaaaaattcaaaattttcgggtttattacagggacagcagagatccagtttggttagtaccgggccctacgcgtccaaggt
cgggcaggaagagggcctatttcccatgattccttcatatttgcatatacgatacaaggctgttagagagataattagaattaatttgactgt
aaacacaaagatattagtacaaaatacgtgacgtagaaagtaataatttcttgggtagtttgcagttttaaaattatgttttaaaatggactat
catatgcttaccgtaacttgaaagtatttcgatttcttggctttatatatcttgtggaaaggacgaaacaccgggtcttcgagaagacacgtttt
agagctagaaatagcaagttaaaataaggctagtccgttatcaacttgaaaaagtggcaccgagtcggtgctttttttggatccaattctac
cgggtaggggaggcgcttttcccaaggcagtctggagcatgcgctttagcagccccgctgggcacttggcgctacacaagtggcctctg
gcctcgcacacattccacatccaccggtaggcgccaaccggctccgttctttggtggccccttcgcgccaccttctactcctcccctagtca
ggaagttcccccccgccccgcagctcgcgtcgtgcaggacgtgacaaatggaagtagcacgtctcactagtctcgtgcagatggacag
caccgctgagcaatggaagcgggtaggcctttggggcagcggccaatagcagctttgctccttcgctttctgggctcagaggctgggaa
ggggtgggtccgggggcgggctcaggggcgggctcaggggcggggcgggcgcccgaaggtcctccggaggcccggcattctgca
cgcttcaaaagcgcacgtctgccgcgctgttctcctcttcctcatctccgggcctttcgacctgcatccatctagatctcgagcagctgaagc
ttaccatgaccgagtacaagcccacggtgcgcctcgccacccgcgacgacgtccccagggccgtacgcaccctcgccgccgcgttcg
ccgactaccccgccacgcgccacaccgtcgatccggaccgccacatcgagcgggtcaccgagctgcaagaactcttcctcacgcgc
gtcgggctcgacatcggcaaggtgtgggtcgcggacgacggcgccgcggtggcggtctggaccacgccggagagcgtcgaagcgg
gggcggtgttcgccgagatcggcccgcgcatggccgagttgagcggttcccggctggccgcgcagcaacagatggaaggcctcctg
gcgccgcaccggcccaaggagcccgcgtggttcctggccaccgtcggcgtctcgcccgaccaccagggcaagggtctgggcagcg
ccgtcgtgctccccggagtggaggcggccgagcgcgccggggtgcccgccttcctggagacctccgcgccccgcaacctccccttcta
cgagcggctcggcttcaccgtcaccgccgacgtcgaggtgcccgaaggaccgcgcacctggtgcatgacccgcaagcccggtgccg
gcggcgggtccggaggagagggcagaggaagtctcctaacatgcggtgacgtggaggagaatcctggcccaatggtgagcaagg
gcgaggaggataactccgccatcatcaaggagttcctgcgcttcaaggtgcacatggagggctccgtgaacggccacgagttcgagat
cgagggcgagggcgagggccgcccctacgagggcacccagaccgccaagctgaaggtgaccaagggtggcccctgcccttcgc
ctgggacatcctgtcccctcagttcatgtacggctccaaggcctacgtgaagcaccccgccgacatccccgactacttgaagctgtccttc
cccgagggcttcaagtgggagcgcgtgatgaacttcgaggacggcggcgtggtgaccgtgacccaggactcctctctgcaggacggc
gagttcatctacaaggtgaagctgcgcggcaccaacttcccctccgacggccccgtaatgcagaagaaaaccatgggctgggaggc
ctcctccgagcggatgtaccccgaggacggcgccctgaagggcgagatcaagcagaggctgaagctgaaggacggcggccacta
cgacgctgaggtcaagaccacctacaaggccaagaagcccgtgcagctgcccggcgcctacaacgtcaacatcaagttggacatca
cctcccacaacgaggactacaccatcgtggaacagtacgaacgcgccgagggccgccactccaccggcggcatggacgagctgta
caagtgagcggccgctaggtacctttaagaccaatgacttacaaggcagctgtagatcttagccactttttaaaagaaaaggggggact
ggaagggctaattcactcccaaagaagtcaagatctgctttttgcctgtactgggtctctctggttagaccagagtctctctggttagaccag
atctgagcctgggagctctctggctaactagggaacccactgcttaagcctcaataaagcttgccttgagtgcttcaagtagtgtgtgcccg
tctgttgtgtgactctggtaactagagatccctcagacccttttagtcagtgtggaaaatctctagcagtttaaacccgctgatcagcctcgac
tgtgccttctagttgccagccatctgttgtttgcccctcccccgtgccttccttgaccctggaaggtgccactcccactgtcctttcctaataaaa
tgaggaaattgcatcgcattgtctgagtaggtgtcattctattctggggggtggggtggggcaggacagcaagggggaggattgggaag
tcaatagcaggcatgctggggatgcggtgggctctatggcggccgttaattaagcttttgttccctttagtgagggttaattgcgcgcttggc
gtaatcatggtcatagctgtttcctgtgtgaaattgttatccgctcacaattccacacaacatacgagccggaagcataaagtgtaaagcct
ggggtgcctaatgagtgagctaactcacattaattgcgttgcgctcactgcccgctttccagtcgggaaacctgtcgtgccagctgcattaa
tgaatcggccaacgcgcggggagaggcggtttgcgtattgggcgctcttccgcttcctcgctcactgactcgctgcgctcggtcgttcggct
gcggcgagcggtatcagctcactcaaaggcggtaatacggttatccacagaatcaggggataacgcaggaaagaacatgtgagca
aaaggccagcaaaaggccaggaaccgtaaaaaggccgcgttgctggcgtttttccataggctccgcccccctgacgagcatcacaa
aaatcgacgctcaagtcagaggtggcgaaacccgacaggactataaagataccaggcgtttccccctggaagctccctcgtgcgctct
cctgttccgaccctgccgcttaccggatacctgtccgcctttctcccttcgggaagcgtggcgctttctcatagctcacgctgtaggtatctca
gttcggtgtaggtcgttcgctccaagctgggctgtgtgcacgaaccccccgttcagcccgaccgctgcgccttatccggtaactatcgtctt
gagtccaacccggtaagacacgacttatcgccactggcagcagccactggtaacaggattagcagagcgaggtatgtaggcggtgct
acagagttcttgaagtggtggcctaactacggctacactagaaggacagtatttggtatctgcgctctgctgaagccagttaccttcggaa
aaagagttggtagctcttgatccggcaaacaaaccaccgctggtagcggtggtttttttgtttgcaagcagcagattacgcgcagaaaaa
aaggatctcaagaagatcctttgatcttttctacggggtctgacgctcagtggaacgaaaactcacgttaagggattttggtcatgagattat
caaaaaggatcttcacctagatccttttaaattaaaaatgaagttttaaatcaatctaaagtatatatgagtaaacttggtctgacagttacca
atgcttaatcagtgaggcacctatctcagcgatctgtctatttcgttcatccatagttgcctgactccccgtcgtgtagataactacgatacgg
gagggcttaccatctggccccagtgctgcaatgataccgcgagacccacgctcaccggctccagatttatcagcaataaaccagccag
ccggaagggccgagcgcagaagtggtcctgcaactttatccgcctccatccagtctattaattgttgccgggaagctagagtaagtagttc
gccagttaatagtttgcgcaacgttgttgccattgctacaggcatcgtggtgtcacgctcgtcgtttggtatggcttcattcagctccggttccc
aacgatcaaggcgagttacatgatcccccatgttgtgcaaaaaagcggttagctccttcggtcctccgatcgttgtcagaagtaagttggc
cgcagtgttatcactcatggttatggcagcactgcataattctcttactgtcatgccatccgtaagatgcttttctgtgactggtgagtactcaac
caagtcattctgagaatagtgtatgcggcgaccgagttgctcttgcccggcgtcaatacgggataataccgcgccacatagcagaacttt
aaaagtgctcatcattggaaaacgttcttcggggcgaaaactctcaaggatcttaccgctgttgagatccagttcgatgtaacccactcgt

gcacccaactgatcttcagcatcttttactttcaccagcgtttctgggtgagcaaaaacaggaaggcaaaatgccgcaaaaaagggaat
aagggcgacacggaaatgttgaatactcatactcttcctttttcaatattattgaagcatttatcagggttattgtctcatgagcggatacatatt
tgaatgtatttagaaaaataaacaaataggggttccgcgcacatttccccgaaaagtgccac

**Fig. S7.** Sequence and map of the plasmids used for DNA methylation editing. More information and the plasmids will be available upon request. Plasmids constructed for this study can be obtained via Addgene (Please check the Addgene website for the final sequence).

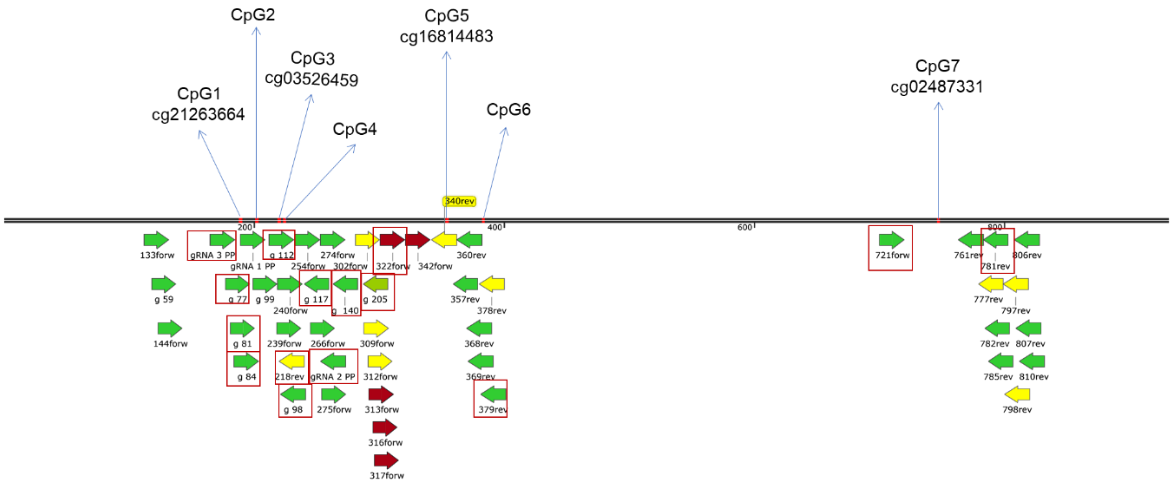


**Fig. S8.** Map of the gRNAs designed to target the 1q21.1 DMR.
All the gRNAs were designed by CRISPOR Version 4.98. The gRNAs in the rectangle were used for the gRNA screening on HEK293T cells (the sequences are listed in the SI Appendix Table 10).
